# Potential Test-Negative Design Study Bias in Outbreak Settings: Application to Ebola vaccination in Democratic Republic of Congo

**DOI:** 10.1101/2020.01.06.20016576

**Authors:** Carl A. B. Pearson, W. John Edmunds, Thomas J. Hladish, Rosalind M. Eggo

**Affiliations:** Department of Infectious Disease Epidemiology & Centre for the Mathematical Modelling of Infectious Diseases, London School of Hygiene & Tropical Medicine; South African Centre for Epidemiological Modelling and Analysis, Stellenbosch University; Department of Biology & Emerging Pathogens Institute, University of Florida

## Abstract

**Background:** Infectious disease outbreaks present unique challenges to study designs for vaccine evaluation. Test-negative design (TND) studies have previously been used to estimate vaccine effectiveness and have been proposed for Ebola virus disease (EVD) vaccines. However, there are key differences in how cases and controls are recruited during outbreaks and pandemics of novel pathogens that have implications for the reliability of effectiveness estimates using this design.

**Methods:** We use a modelling approach to quantify TND bias for a prophylactic vaccine under varying study and epidemiological scenarios. Our model accounts for heterogeneity in vaccine distribution and for two potential routes to testing and recruitment into the study: self-reporting and contact-tracing. We derive conventional and hybrid TND estimators for this model and suggest ways to translate public health response data into the parameters of the model.

**Results:** Using a conventional TND study, our model finds biases in vaccine effectiveness estimates. Bias arises due to differential recruitment from self-reporting and contact-tracing, and due to clustering of vaccination. We estimate the degree of bias when recruitment route is not available, and propose a study design to eliminate the bias if recruitment route is recorded.

**Conclusions:** Hybrid TND studies can resolve the design bias with conventional TND studies applied to outbreak and pandemic response testing data, if those efforts collect individuals’ routes to testing. Without route to testing, other epidemiological data will be required to estimate the magnitude of potential bias in a conventional TND study. Since these studies may need to be conducted retrospectively, public health responses should obtain these data, and generic protocols for outbreak and pandemic response studies should emphasize the need to record routes to testing.

**Key Messages:**

- Conventional Test-Negative Design (TND) studies can be biased when follow-up of cases leads to testing and recruitment, which occurs for example during contact tracing.
- A hybrid TND estimator can eliminate this bias, if individual testing routes are recorded.
- The related bias in the conventional TND estimator can be quantified using epidemiological measures.
- If testing route data is unavailable, bias can be limited by other study measures.

## Background

Study designs to evaluate new vaccines during outbreaks and pandemics are challenging for logistical, epidemiological, social, and ethical reasons (1-7). Novel and poorly understood pathogens can rapidly overwhelm local health systems, subsequently enabling other crises, or require intensive control policies, complicating both response efforts and research (8-11). However, some key pathogens are only routinely observable under these conditions, like Ebola virus disease (EVD) and others on the World Health Organization (WHO) R&D Blueprint priority list (12).

During outbreaks of highly pathogenic infections there may be pressure to introduce experimental vaccines as quickly as possible (13, 14), as well as resistance to classical randomised controlled trials (15). For EVD, the existence of a licensed vaccine (rVSV-ZEBOV) (16-19) further complicates trials for new vaccines. Such circumstances suggest alternative evaluation strategies, and a test-negative design (TND) study has been proposed to evaluate a two-dose vaccine in eastern Democratic Republic of Congo (DRC) (20-22). This design estimates vaccine effectiveness from the odds ratio for test outcome conditional on vaccination status and has lower misclassification bias than traditional case-control studies (23, 24).

TND studies have been used to estimate the effectiveness of vaccines against influenza (25), rotavirus (26-28), pneumococcus (29), and other pathogens (30, 31). The approach can also assess interventions such as vector control and risk-factor management (32-34). TND studies recruit people with symptoms, test those recruits using a highly sensitive and specific method to separate cases (test-positives) from controls (test-negatives), and finally sort them by vaccination status (35). TND studies can be retrospective, potentially using stratification by other factors to limit confounding effects. For influenza, TND studies usually recruit people seeking care for influenza-like illness, ascertain vaccine status by self-report, and determine infection status by RT-PCR, though specifics vary (25, 30).

To obtain unbiased estimates, the following criteria must be met: i) transmission occurs in a population with partial vaccine coverage; ii) vaccination status does not affect symptom rates unrelated to the target pathogen; iii) given symptoms in an individual, care-seeking behaviour does not vary by underlying cause; iv) an individual’s past recruitment as a control (even multiple times) must not prevent subsequent recruitment as a case; and v) there is no misclassification of individuals’ infection or vaccine status (33, 36).

Here we examine how outbreaks present novel misclassification problems for TND studies and how this can bias TND vaccine effectiveness estimates, and we quantify how that bias varies with differences in vaccine distribution, recruitment, risk of infection, and testing practice. We introduce a hybrid design that can address these biases, and we identify alternative steps to mitigate potential bias.

## Methods

### Key Differences During Outbreaks

During outbreaks and pandemics, testing is used both to make treatment decisions for individuals and to trigger public health responses (*e*.*g*. testing conducted post-mortem or of asymptomatic contacts). For example, during EVD outbreaks, suspect cases may be tested because of i) presence of symptoms or ii) high-risk contact with a known case (37, 38). We represent these testing reasons as distinct recruiting routes: self-reporting people who seek care for EVD-like symptoms (analogous to influenza studies); and people identified via active contact-tracing from a confirmed case (commonly part of public health control efforts during outbreak and pandemic responses). However, depending on how data are collected, we may know total numbers for each testing route, but not matched to outcome. For clarity, we provide specific definitions for our model terms (Table 1).

**Table 1:** Definitions of terms used in this analysis.

| Term | Definition |
| --- | --- |
| **Recruitable population** | The total population who may later be *recruited* into the *study*. In practice, unknown when the *vaccination campaign* occurs, but known in the model. |
| **Vaccination campaign** | Administration of the *study vaccine* to some of the *recruitable population*. |
| **Study** | A TND study of the *study vaccine* against EVD in the *recruitable population*. |
| **Targeted** | The portion of the *recruitable population* who could be vaccinated, with vaccination homogeneously distributed. *Non-targeted* refers to the complementary portion of the *recruitable population*: none of these individuals receive the vaccine. |
| **Recruitment** | Testing for potential EVD infection and being counted in the *study*; distinct from being *targeted* (for *vaccination*). Occurs via two routes: *self-reporting* and *contact-tracing*. |
| **Self-reporting (SR)** | Testing of individuals without a known link to a previous case. |
| **Contact-tracing (CT)** | Testing of individuals because of contact with a confirmed EVD case. |
| **N, U, and V** | Populations for non-targeted (N), or targeted and unvaccinated (U) or vaccinated (V). |
| **Annotation ‘ and “** | Recruitment route indicators; e.g. V’ would be vaccinated individuals recruited via self-reporting vs N” would be non-targeted individuals recruited via contact-tracing. |
| **Subscript + and -** | Test outcome indicators; e.g. U’<sub>-</sub> would be unvaccinated individuals, recruited via self-reporting, that test negative vs V”<sub>+</sub> would be vaccinated individuals, recruited via contact-tracing, that test positive. |

### Vaccination Model

Vaccinated individuals avoid infection if exposed to EVD with probability *E*, the true vaccine efficacy. We assume all-or-nothing vaccine protection. The study measures *Ê*, the estimated vaccine effectiveness. While there may be a delay from vaccination to protection, *e*.*g*. a multiple dose requirement, we model a scenario where protection has already occurred in vaccinated individuals. Aside from preventing EVD, we assume vaccination has no effect.

We represent vaccine distribution by dividing the recruitable population into two types of individuals: those targeted by the vaccination campaign and those not (Figure 1), referred to as the targeted, *p*_*in*_, and non-targeted, 1-*p*_in_, fractions (Table 1 and Table 2). Among the targeted population, only some individuals receive the vaccine, leading to a coverage level, *L*, in that population. Because we consider a situation where there is already a licensed, efficacious vaccine, we assume there is no distribution bias (*e*.*g*. prioritization of healthcare workers) within the targeted population. We assume individuals cluster by targeted status, such as might occur if study vaccine distribution targeted particular villages, so the contacts of self-reporting cases always have the same target/non-target status as the associated case. Aside from these distinctions, all individuals are identical.

**Table 2:** Parameter Summary. This table summarizes the measurements and model parameters used in this analysis. We also introduce an alternative parameterization of the recruitment model, which is less intuitive when describing the model but more useful for understanding the impact on bias.

| Vaccination Parameters |  |  |  |
| --- | --- | --- | --- |
| Symbol | Name | Description | Calculation |
| $E$ | True vaccine efficacy | The probability of preventing disease given an EVD exposure | Estimation target |
| $\hat{E}$ | Estimated vaccine effectiveness | | Estimator, Equations 1-3 |
| $p_{in}$ | targeted fraction | The fraction of the recruitable population with some vaccine coverage; the *non-targeted fraction*, $1-p_{in}$, has no vaccine coverage | *see Supplement Section S7* |
| $L$ | vaccine coverage | In the targeted population, the achieved vaccine coverage | *see Supplement Section S7* |
| Outbreak Response Metrics |  |  |  |
| $SR_+$ | Self-reported test-positive | Total number of individuals who test positive when they self-report to a health centre | Estimated from outbreak response metrics if available, either before or after start of vaccine campaign |
| $SR_-$ | Self-reported test-negative | Number who test negative when they self-report to a health centre | |
| $CT_+$ | Contact-traced test-positive | Number who test positive after identification by contact-tracing from a known case | |
| $CT_-$ | Contact-traced test-negative | Number who test negative after identification by contact-tracing from a known case | |
| Recruitment Parameters |  |  |  |
| $B$ | Self-reporting test-negative rate | The expected number of self-reporting test-negatives per self-reporting test-positive case | $\frac{SR_-}{SR_+}$ |
| $\lambda$ | Contact-tracing test rate | The expected number of tested contacts per self-reporting test-positive case | $\frac{CT_+ + CT_-}{SR_+}$ |
| $R''$ | Contact-tracing test-positive rate | The expected number of new infections among tested contacts of a known case, when the study vaccine is not present, per self-reporting test-positive case | $\frac{CT_+}{SR_+}$ |
| Alternative Recruitment Parameters |  |  |  |
| $f_-$ | Self-reporting test-negative fraction | The proportion of self-reporting individuals that test-negative | $\frac{B}{B+1}$ or $\frac{SR_-}{SR_+ + SR_-}$ |
| $\rho$ | Recruitment route ratio | The ratio of contact-tracing recruitment to self-reporting recruitment | $\frac{\lambda}{B+1}$ or $\frac{CT_+ + CT_-}{SR_+ + SR_-}$ |
| $p_t$ | Contact-tracing test-positive fraction | The test-positive fraction of direct contact-tracing recruitment in the absence of vaccination | $\frac{R''}{\lambda}$ or $\frac{CT_+}{CT_+ + CT_-}$ |

**Figure 1.**
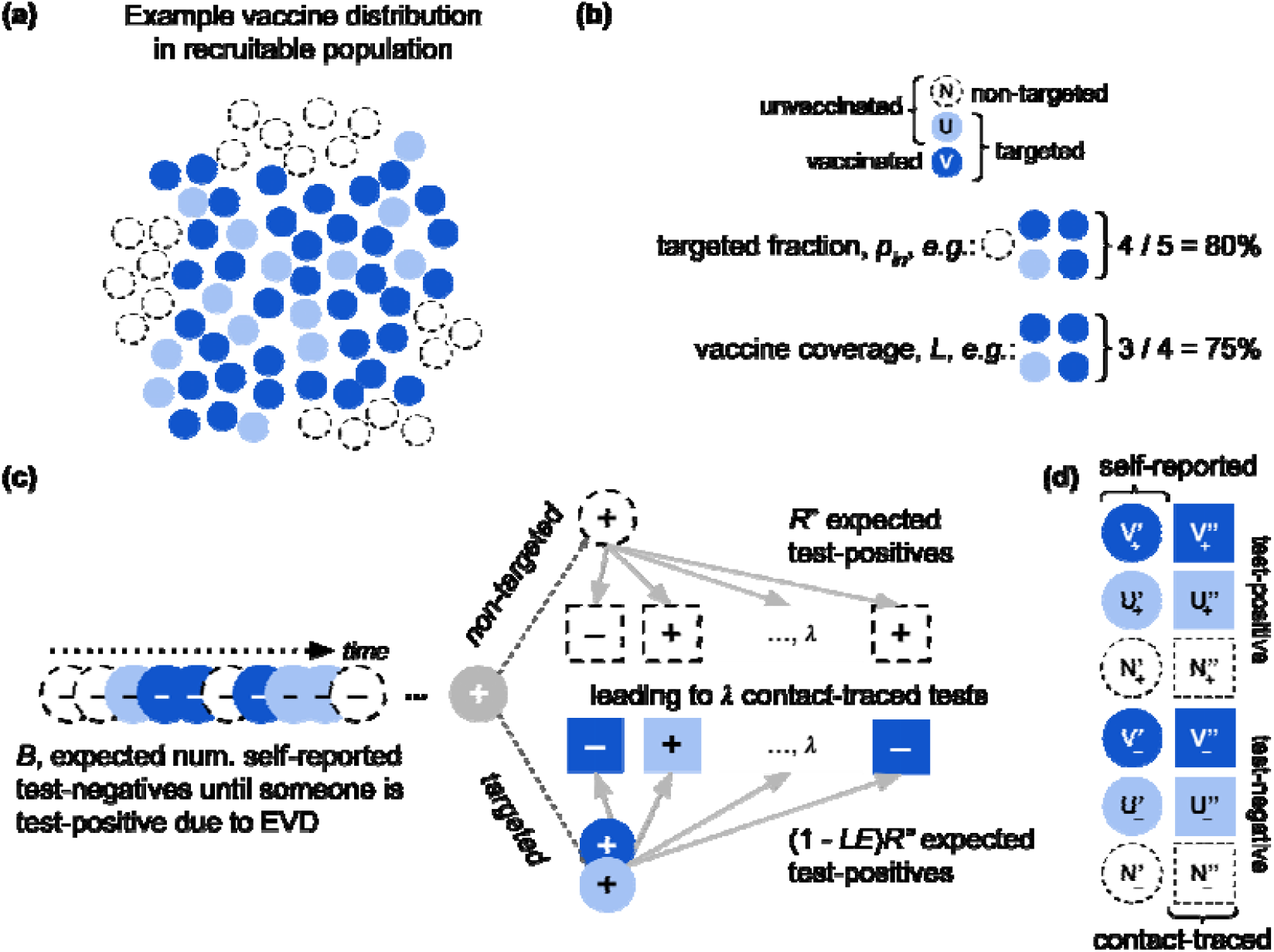
The modelled population and recruitment into the TND study. (a) Individuals and their contacts are either targeted for vaccination (filled circles – dark blue receive the vaccine and light blue do not) or not (open circles). (b) The fraction that are targeted (and thus may be vaccinated) is p_in_; none of the non-targeted population (open circles, *N* label) receives the vaccine. Of those targeted, some are not vaccinated (e.g. because they are ineligible due to age, pregnancy, recent illness, immunocompromised status; or because there is only sufficient study vaccine to deliver partial coverage) (light blue, *U* label) and some are (dark blue, *V* label). The vaccine coverage in the targeted population is *L*. In the recruitable population (the combination of *N, U*, and *V*), non-vaccinees (*N* and *U*) are infected on EVD exposure, while vaccines (*V*) avoid disease at the vaccine efficacy, *E*. (c) An expected number of self-reported people test negative (circles with – sign), *B*, until a test-positive (circle with + sign) is identified. This leads to an expected amount of follow-up testing, λ, which finds R” more cases if the initial case is in the non-targeted population, and (1-LE)R” if targeted. The coverage, *L*, efficacy, *E*, and targeted fraction, p_in_, determine the likelihood of observing the self-reporting case among targeted vs non-targeted individuals and vaccinated vs unvaccinated individuals. (d) Resulting categories that can be recruited into the study. *U* and *V* are, respectively, the unvaccinated and vaccinated individuals in the targeted population; N are non-targeted individuals. The ‘vs “annotations indicate, respectively, self-reporting vs contact-traced, and the – vs + subscripts indicate test-negative and test-positive outcomes, respectively.

Ideally, a study would recruit only from targeted populations, but these may not be distinguishable in practice. Indeed, in reality there may be many distinct populations, for example areas with different vaccination coverage; we consider just two to focus on the impact of heterogeneity.

### TND Recruitment Model

We identify recruits by their testing route, either people who self-report or are contact-traced from a confirmed case (Table 1 and Figure 1), because we assume these have different criteria for testing (*i*.*e*. in our application, the self-reporting criteria are more stringent). In the Supplement, we generalise these as *primary* and *secondary* recruitment routes. We assume that introduced cases in the recruitable population are found before any contacts have become symptomatic and therefore infectious (though they may be infected but not yet symptomatic), consistent with the typical experience in the rVSV-ZEBOV trial in Guinea and deployment in DRC (17, 39). In those efforts, after identifying an index case via self-reporting, the response programmes contact-traced around that individual (an index case) to identify potential exposures (direct contacts) and to identify people for vaccination (contacts-of-contacts) under the ring protocol. This assumption means that in the model all self-reported cases result from external introductions, and all contact-traced recruits are only exposed to a single case.

Consistent with the WHO guidance for EVD outbreak response (38), we assume self-reporting individuals present with multiple EVD-like symptoms. Both EVD and other causes of EVD-like symptoms lead to these self-reports, resulting in test-positive and test-negative outcomes, respectively. Targeted and non-targeted populations are assumed to have equal rates of EVD exposure, but on average the targeted population has fewer EVD cases due to the vaccine. Other causes of EVD-like symptoms occur at the same rate in both populations. During an outbreak, rates of EVD exposure and of other causes of EVD-like symptoms may vary, but our analysis only depends on a consistent, long-term average relative rate of these processes. As such, we can define parameters as per self-reported test-positive, and we use the background self-reporting test-negatives per test-positive rate, *B*, to represent the self-reporting process overall (Table 2).

We also represent testing of traced contacts based on the outbreak response control protocols, which direct testing based on high-risk interaction and fever (38). However, fever is common in EVD-prone areas and may be subjectively evaluated, so we assume that this criterion is practically based on high-risk contact. Thus, the average number of tested contacts is λ, which we assume is the same irrespective of targeted or vaccination status (Supplement Section S3.3 relaxes this assumption). These contacts are exposed or not, according to the contact-tracing test-positive rate, *R”*, which is reduced within the targeted population by the vaccine.

### Translating outbreak metrics to estimate bias

To evaluate a particular study’s potential bias, we need real-world outbreak response metrics to estimate model parameters. For studies augmenting an ongoing outbreak response, data already collected could be used. For example, partial data on the number of tested individuals, stratified by test outcome and testing route could be used to bound model parameters (Table 2).

The model also depends on how the study vaccination is distributed, represented by targeted fraction and coverage within that fraction, *p*_in_ and *L* (respectively). Depending on the study protocol, these could be ascertained in different ways (Supplement Section S7).

### Summary of assumptions

- The study period and population are sufficient to use expected values and minimize the impact of heterogeneity, for example superspreading events.
- Cases and their contacts have the same targeted status.
- All individuals have the same exposure risk to EVD and other causes of EVD-like symptoms, average number of contacts, and risk of infection per contact.
- Non-vaccination among targeted populations happens randomly.
- There are different testing criteria for self-reporting and contact-tracing individuals.
- Self-reporting cases are identified before anyone they have infected becomes symptomatic, and contact tracing prevents transmission among contacts.

### TND Estimator for Outbreak Context

There are twelve recruitment categories in our model, based on targeted and vaccination status (N, U, and V for non-target, unvaccinated, and vaccinated respectively), test outcome (subscripts – and +), and testing route (annotations ‘and “for self-reporting and contact-tracing respectively) (Table 1 and Figure 1). The conventional TND estimator:

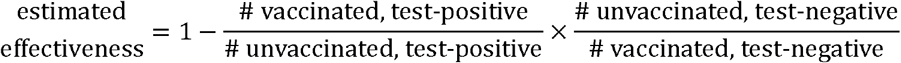

can be written with these categories as:

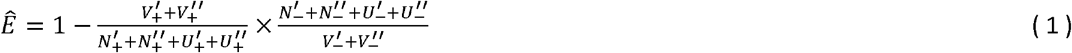

The expected counts of these categories can be expressed with the six model parameters we defined (full derivation in Supplement S3-S4). Recall these are true efficacy (*E*), the targeted fraction of the recruitable population (*p*_in_), the vaccine coverage in targeted population (*L*), the number of self-reporting test-negatives per test-positive (*B*), average number of contacts tested per self-reporting test-positive (λ), and average number of contacts that test-positive (*R”*). Substituting these, we can obtain:

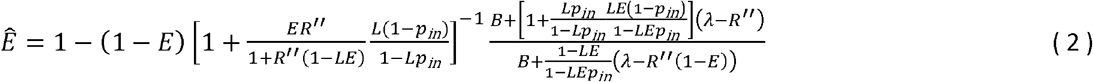

The terms to the right only cancel under very specific circumstances, thus the bias is generally non-zero and the magnitude is a function of all model parameters. We refactor Eq. (2) with alternative parameters relating to recruitment and epidemiological measures from the outbreak, namely the fraction of tests that are negative for self-reporting individuals (*f*_–_), the ratio of contact-traced to self-reporting individuals (ρ), and the fraction of tests that are positive among contact-traced individuals in the absence of vaccination (*p*_t_) (Supplement Section S4). We use this form to explore the bias and to evaluate potential maximum bias under specific outbreak scenarios:

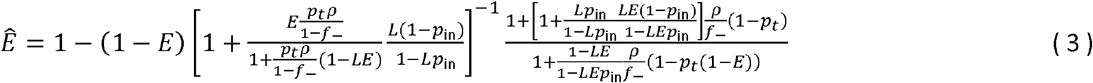

We also consider an alternative study design, where testing route is always available. In this design, we combine the conventional TND estimator with a conventional cohort study estimator. We evaluate the self-reporting individuals with the TND component and the contact-traced individuals with cohort estimator:

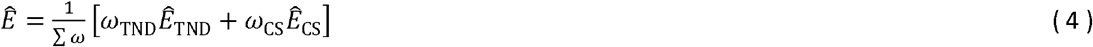

### TND Estimator Bias

Across a wide range of self-reporting test-negative fractions (*f*_-_), contact-tracing test-positive fractions (*p*_*t*_), targeted fractions (*p*_*in*_), and testing route ratios (*ρ*), the absolute error in *Ê* is ≤ 0.1(Figure 3). If information from the outbreak response indicates these parameter values are bounding, then our model indicates conventional study bias lies in that range. Ideally, individual testing routes will be available, and the hybrid design can be used. When the hybrid design cannot be used, we have identified two avenues to limit bias, restricting recruitment to either targeted or self-reporting individuals only.

#### Hybrid Design: No Recruitment Bias

If study data permit stratifying recruits by testing route, then an alternative study design can be used to eliminate the design bias (Figure 2). Consider a cohort study-like estimator of vaccine effectiveness using only the contact-traced recruits:

**Figure 2.**
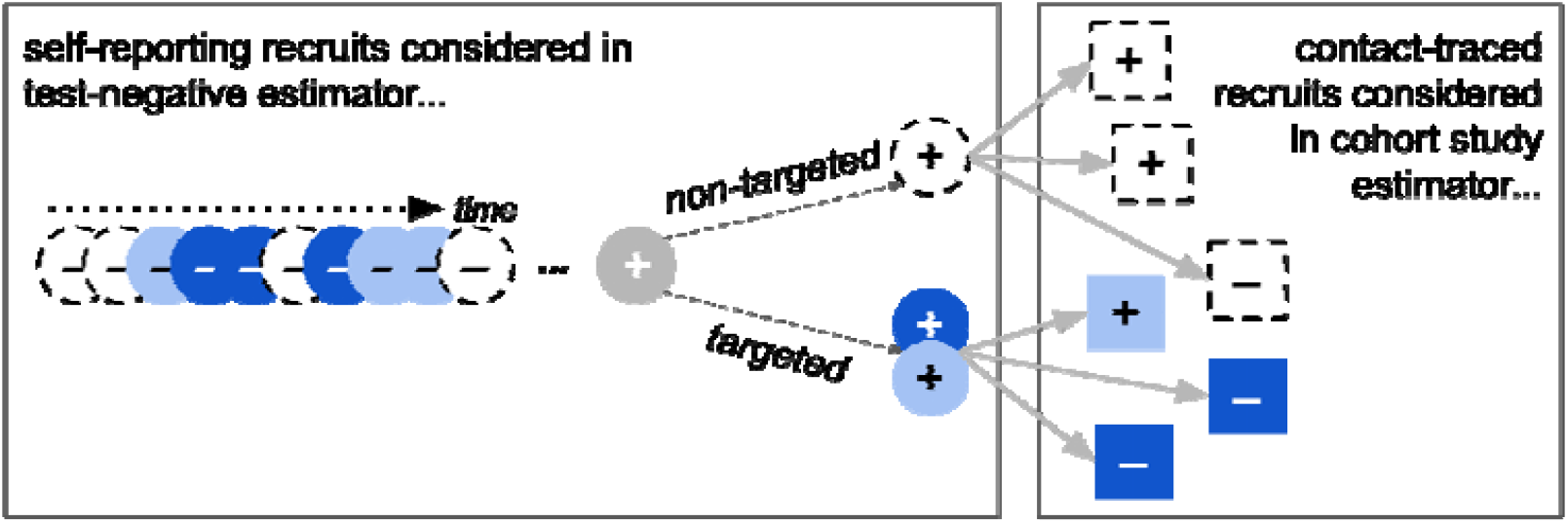
Recruitment into Hybrid Study. An alternative study design combining the test-negative estimator for self-reporting individuals and a cohort-study style estimator for contact-traced individuals. This arrangement eliminates the design bias identified for the conventional TND but requires that individuals be stratified by recruitment route.

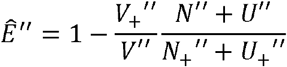

Under our model assumptions, this estimator is asymptotically equal to the true efficacy, *E*. The positive fraction of contact-traced individuals will be a context specific transmission probability. For vaccinees, it is reduced by (1-*E*). The transmission-related term cancels from the two fractions, leaving only *Ê* = 1 – (1 – *E*) = *E*, indicating this estimator is asymptotically unbiased. When only considering self-reporting individuals, our model assumptions match the TND requirements for an unbiased estimate, and therefore the test-negative estimator limited to self-reporting individuals is also asymptotically unbiased. Thus, any weighted combination of the two estimators like Eq. (4) is also asymptotically unbiased.

#### Restricting Recruitment to Targeted Populations

Ideally, a study would strictly recruit from populations that were targeted for vaccination. This could be achieved either by expanding the targeted (for vaccination) population to coincide with the potentially recruitable (by testing) population, or by censoring the tested population to only the population targeted for vaccination. The former may be possible with, for example, an extensive community engagement programme that results in homogenous coverage across a wide region. The latter may be possible if there is additional data collected, like place of residence, that allows targeted population, *i*.*e*. *p*_*in*_ = 1, some bias remains due to recruitment of contact-traced exclusion of individuals outside the targeted population. Even if the study is constrained to only a individuals, but it no longer depends on the vaccine coverage (see Supplement Section S5.1). However, uniformly distributing a vaccine among the population will be complicated in an outbreak response setting, and the bias is sensitive to other factors even when most of the recruited population was targeted (Figure 4).

**Figure 3:**
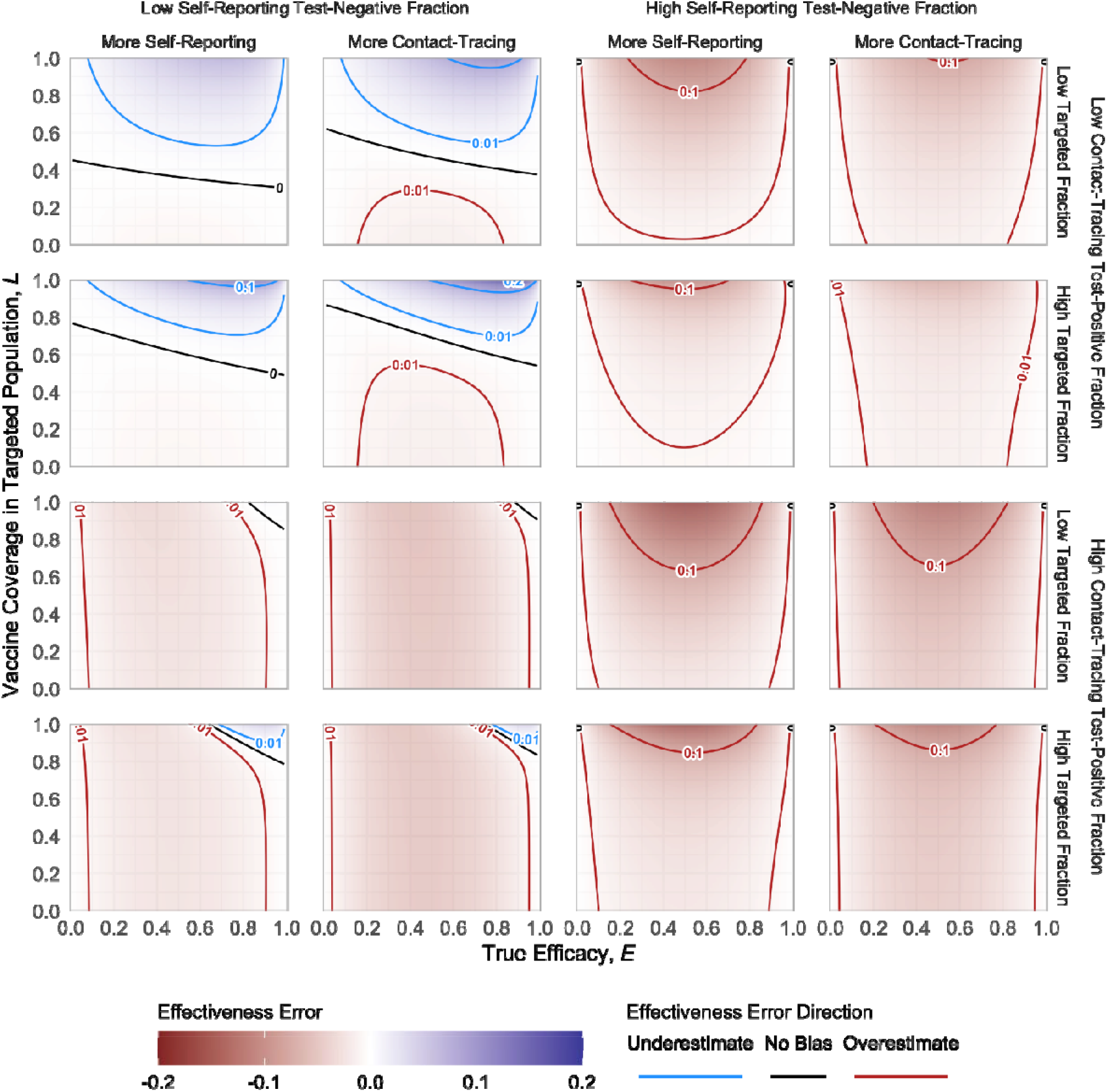
Bias Trends Across All Model Parameters. The figure illustrates the bias trends with respect to true efficacy, *E*, and vaccination coverage in the targeted population, *L*. The sixteen panels correspond to combinations of example values for: (outer columns) self-reporting test-negative fraction (at low = 0.8 and high = 0.99); (inner columns) the recruitment route ratio (ρ at low = 0.5 and high = 2; less than 1 implies more self-reporting recruitment, greater than 1 implies more contact-tracing recruitment); (outer rows) contact-tracing test-positive fraction (at low = 0.1 and high = 0.3); and the targeted fraction (p_in_, at low = 0.6 and high = 0.9).

**Figure 4:**
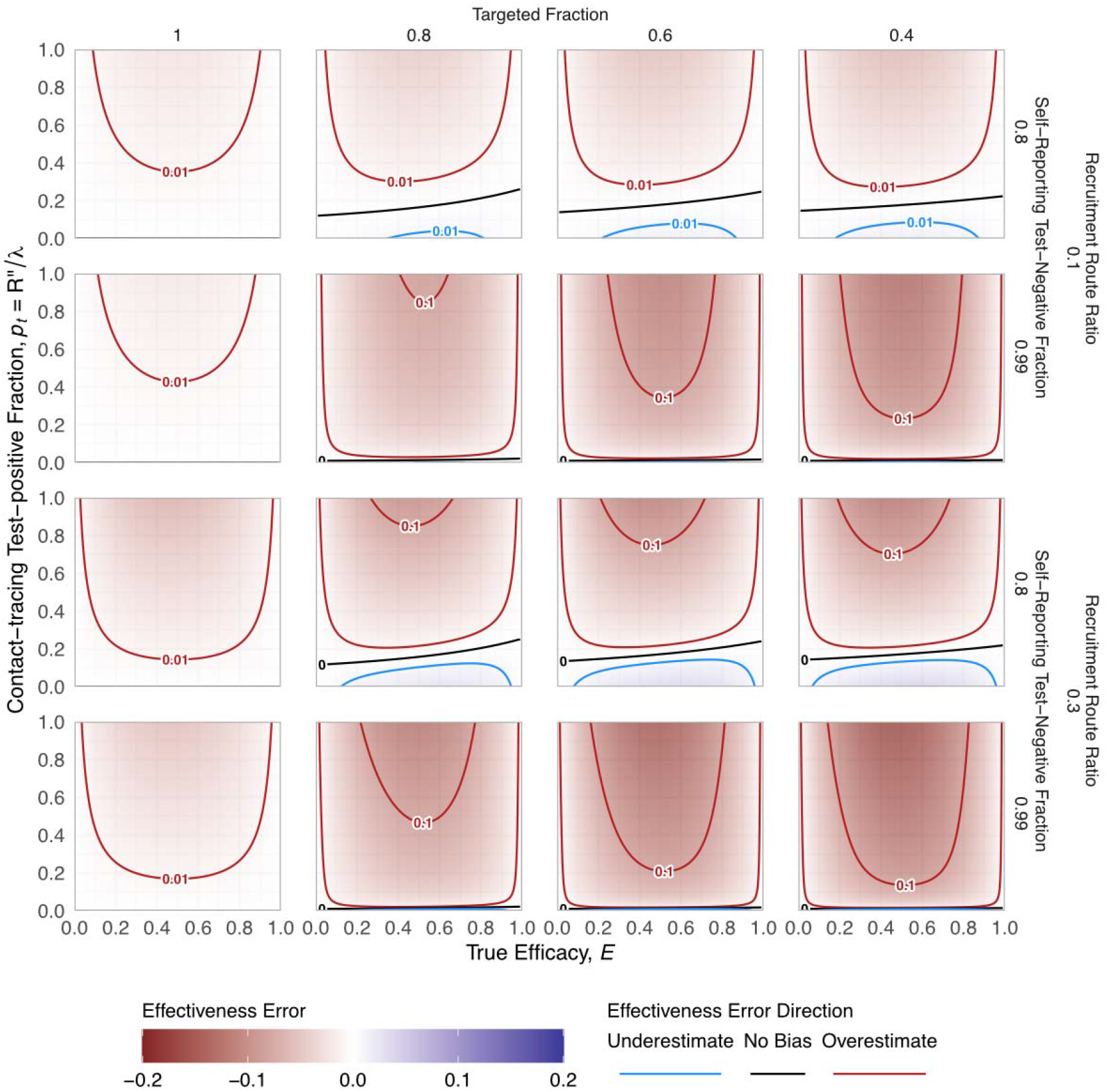
Impact of Decreasing Targeted Fraction Among Recruits. The panels show decreasing targeted fraction (columns from left to right) for scenarios stratified by self-reported test-negative fraction in recruitment (0.8 and 0.99) and recruitment route ratio (0.1 and 0.3) (rows). This figure shows 70% coverage level in the targeted population, *L*=0.7.

If the targeted fraction decreases, generally the magnitude of bias in the estimate increases. Bias generally peaks when the true vaccine efficacy is around 50% and goes to zero as true efficacy approaches 0 or 100%. Because initial cases are more likely among non-targeted populations, contact-traced individuals are likewise biased towards being non-targeted and thus un-vaccinated. Thus, bias tends towards overestimation as contact-traced individuals more frequently test positive. This can reverse for high levels of contact-traced recruitment, when most contact-traced individuals are test-negative. All other factors being equal, more coverage means more extreme bias as targeted and non-targeted populations diverge.

#### Restricting Recruitment to Self-Reported individuals

The bias can also be corrected by restricting recruitment strictly to self-reported individuals. If perfectly achieved, then the bias is 0 (see Supplement S4.4). During outbreak response, priorities and data processes, however, may focus on identified cases, neglecting detailed tracking of test-negatives. Thus, in retrospective analyses, test-negative data might only reflect vaccine status, not route to testing, while more detailed data for test-positives is available. If that applies, the resulting estimator bias is:

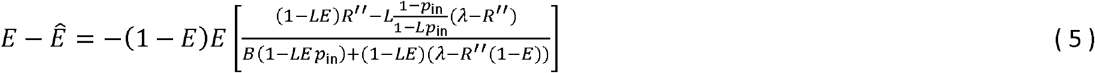

Minimising, and ultimately eliminating, bias in Eq. (5) still depends on maximising *p*_in_. One way of achieving high *p*_in_ is excluding non-targeted populations. This suggests a trade-off between precision and bias: if the study invests the effort to exclude any non-targeted population it may make sense to include contact-traced recruits.

If contact-traced test-positives are excluded retrospectively, the resulting bias magnitude may be lower even if contact-traced test-negatives are included due to misclassification (Figure 4 vs Figure 5). However, the direction of bias changes with changing targeted fraction (*p*_in_): the no-bias line falls at higher contact-tracing test-positive fractions (*p*_t_) when targeted fraction decreases. The magnitude of bias at the extremes of the contact-tracing test-positive fractions is driven largely by the number of self-reporting test-negatives. Other factors being equal, fewer self-reported test-negatives means a lower self-reporting test-negative fraction (*f*_–_ = 0.8 versus 0.99) and higher testing route ratio (ρ = 0.3 versus 0.1), both of which correspond to more extreme bias.

**Figure 5:**
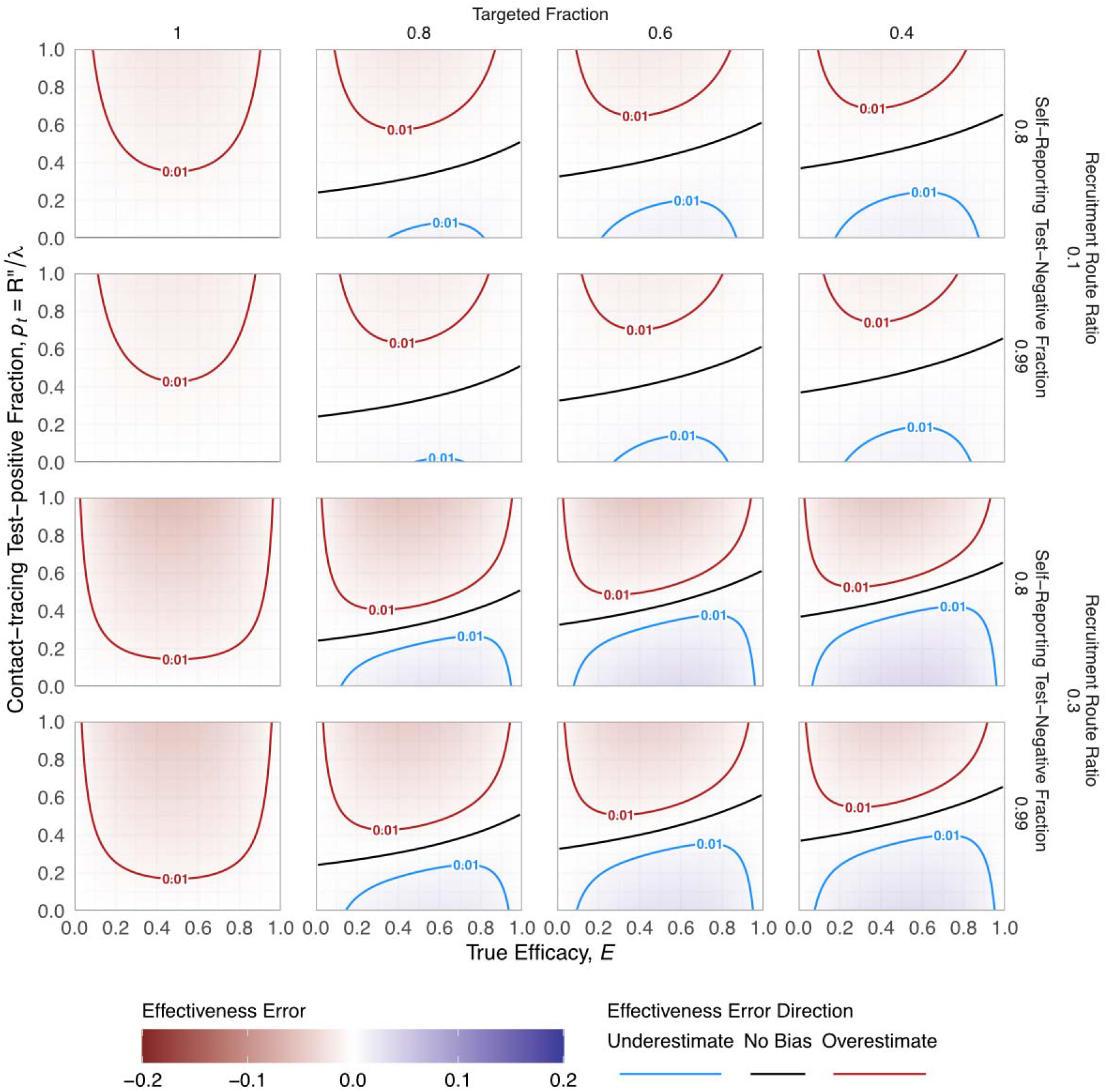
Bias Due to Inability to Exclude Contact-Traced Test-Negatives. The panels show decreasing targeted fraction (columns from left to right) for scenarios stratified by self-reported test-negative fraction in recruitment (0.8 and 0.99) and recruitment route ratio (0.1 and 0.3) (rows). This figure shows 70% coverage level among targeted individuals, *L*=0.7. The range of bias is usually smaller than when recruitment is restricted to the targeted population only (Figure 4).

#### Quantifying Potential Bias from Outbreak Response Metrics

To quantify the bias range for an EVD vaccine study in DRC, we determined the plausible range of outbreak response metrics and corresponding model parameters (Table 2). We consider test positive counts SR_+_ in (100, 150) and CT_+_ in (100, 400), for self-reporting and contact-tracing respectively. We consider test negative counts SR- in (6500, 7000) and CT- in (900, 1200), for self-reporting and contact-tracing respectively.

When restricting recruitment to targeted populations only, the bias in estimated effectiveness is less than 3% overestimation (Figure 6, left panel), but can increase to >15% overestimate if coverage is high and targeted fraction is low (Figure 6, right panel; 90% coverage, 40% targeted fraction). As a larger non-targeted population is recruited, increasing coverage increases bias, corresponding to the increasing distinction in infection risk between targeted and non-targeted populations. For these outbreak response metrics, the estimate of vaccine effectiveness consistently exceeds the true efficacy.

**Figure 6:**
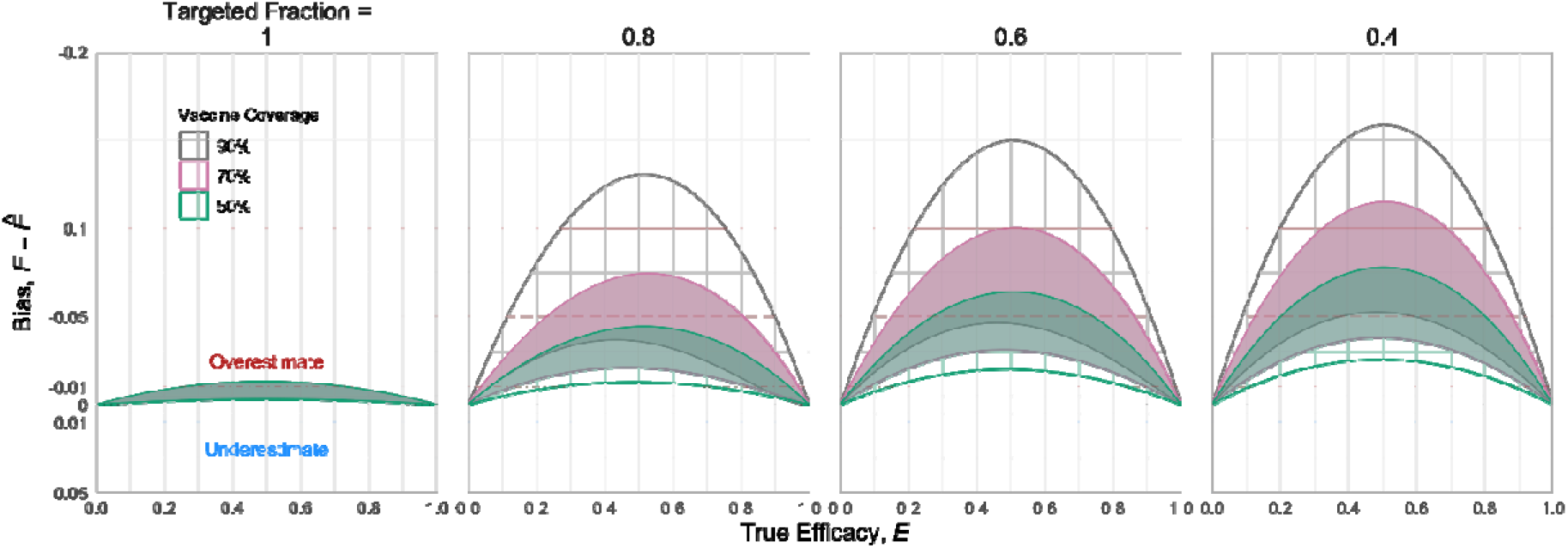
Bias possible when recruiting targeted individuals only. These bias envelopes were computed assuming outbreak response metrics, , , and, which corresponds to 97.7-98.6% of self-reporting cases testing negative, testing 6-16 contact-traced individuals per self-reported case, and 10-25% of those contact-traced individuals testing positive. If the study is restricted to recruit only the targeted population (left-most panel), then bias can be limited to less than 3% overestimation. However, as the targeted fraction falls, the error range generally increases, to >15% peak bias for high coverage (90%) and low targeted fraction (40%). Higher coverage in the targeted population generally increases bias; this reflects increasing differences between the targeted and non-targeted individuals.

When restricting recruitment to self-reported individuals only, with no misclassification of testing route for test-negatives, there is no bias (Figure 7, left panel). As the misclassification increases from 0 to 100%, the magnitude of bias increases and tends towards underestimation, though the range of possible bias includes overestimation. For the most extreme case, where all contact-traced test-negatives are included in a scenario with a low targeted fraction and high coverage, the bias spans roughly 1% overestimate to 5% underestimate (Figure 7, right panel; 90% coverage, 40% targeted fraction). As with restricting recruitment to the targeted population, the magnitude of the bias increases with coverage among targeted individuals.

**Figure 7:**
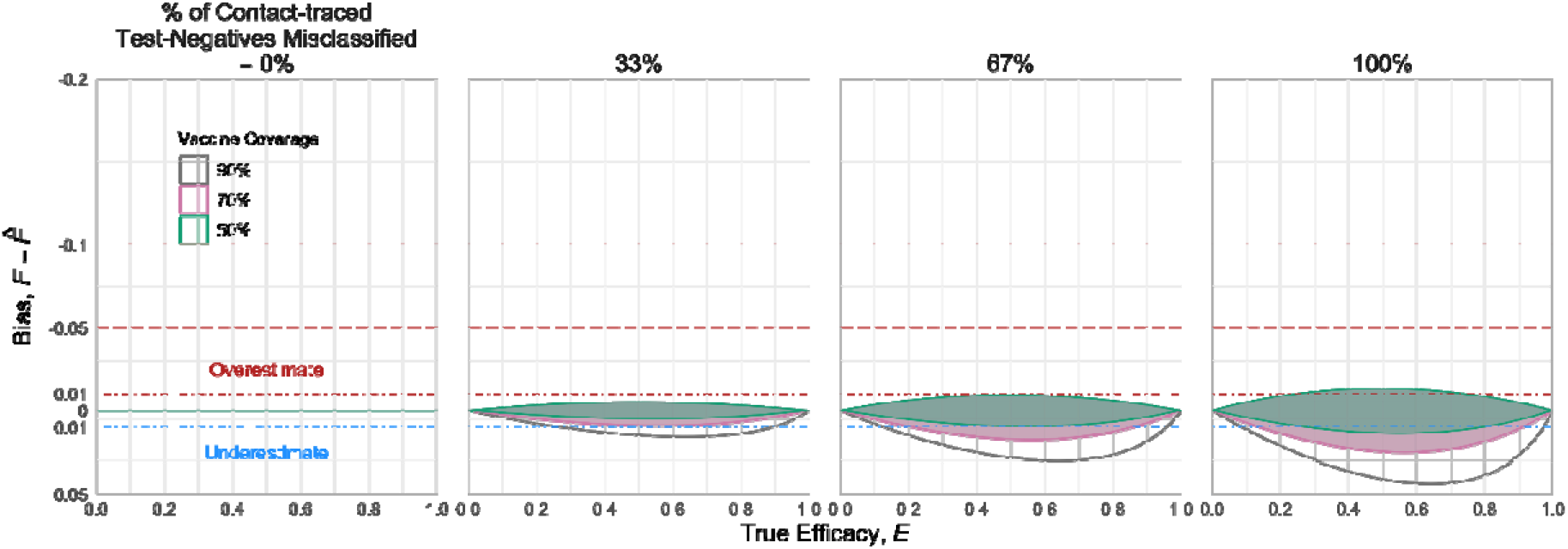
Bias possible when recruitment is restricted to self-reported individuals only. If the study analysis is able to restrict recruits to only self-reporting individuals, then there is no bias (left panel). However, as contact-traced test-negative individuals are increasingly included (moving right across panels), bias range increases to between 1% overestimate and 5% underestimate. However, this range is notably smaller than if only recruiting from the targeted population (Figure 6). As with restricting recruitment to targeted individuals only, higher levels of coverage lead to wider bias range. These ranges reflect the same parameters used in Figure 6, including targeted fraction. In this figure, the overestimate bounds (upper ribbon lines) closely align.

## Discussion

Previous work has explored biases in TND studies due to care-seeking or other confounding and selection effects (35, 40, 41), test or vaccine status misclassification errors (25, 42-44), and vaccine mechanism (36, 45). Here, we demonstrate that public health response efforts can also generate bias in the effectiveness estimates of a conventional TND study. If the response effort collects data on route to testing, a hybrid design study is possible and does not suffer this bias. Retaining this information should therefore be a high priority. This hybrid design warrants further analysis, as we have only considered it with a simplified model.

When a conventional TND study cannot distinguish self-reported and contact-traced recruits, or whether recruits were generally among a population targeted for vaccination, then the vaccine effectiveness estimate is potentially biased. These are both real, practical problems: in addition to general difficulty of collecting data during an outbreak, it may be difficult to achieve uniform levels of coverage when deploying a vaccine in an emergency setting, particularly in highly mobile populations or those affected by civil unrest.

In our model, the bias arises from the interaction of heterogeneous vaccination distribution and the inclusion of tested individuals from contact-tracing. Because initial cases found through self-reporting are more likely to be non-targeted (and thus unvaccinated) individuals, including contact-traced recruits overrepresents those individuals in the estimator. This in turn can tilt the bias either towards or away from the null, depending on how infection risk and testing criteria differ in the contact-traced population versus the general population. If the self-reporting process leads to many more test-negative recruits than recruits from contact-tracing (either positive or negative), then the bias from contact-tracing is relatively smaller. If it does not, then the relative number of cases versus controls from contact-tracing will determine the general direction of bias: more cases lead to overestimation, more controls lead to underestimation.

While the ideal solution is to maintain data on route to testing, if that does not occur we show that the range of potential bias can be quantified with aggregate epidemiological data from an outbreak. For the range of outbreak response metrics we used to represent the EVD outbreak in DRC, this is less than 10% if the study can achieve high targeted fraction (*p*_in_ ≥ 0.6) with at most moderate coverage (*L* ≥ 0.7).

Practically, it may be possible to limit but not eliminate these drivers of the bias. For the EVD epidemic in eastern DRC, responders tried to test individuals meeting one of the outbreak “suspected case” definitions, which combine different levels of symptoms and potential contact with known cases (37, 38). This practice would likely continue in populations that received a study vaccine, meaning this testing process would be the likely source for a retrospective TND study of a new vaccine.

We framed our analysis in terms of event counts, but it can also be thought of in terms of testing thresholds. For example, we frame contact-tracing recruitment as a number of contacts and the number of cases among them. The resulting effectiveness estimator error is then driven by the ratio of those values. In an infectious disease sense, this ratio is the transmission probability, but it could instead be interpreted in terms of the sensitivity of the criteria for testing: should testing criteria be stricter to conserve limited resources, or more relaxed to ensure no positives are missed? A similar analogy applies for testing self-reporting individuals. For both self-reported and contact-traced testing, bias in the vaccine effectiveness estimate is generally lower when testing criteria are less strict (i.e. the protocol is more sensitive), consistent with a control-oriented outbreak response, while a more resource-limited response would have higher bias.

Our analysis describes vaccination during EVD outbreaks, but our work has general implications for evaluating interventions in other infectious disease settings with a public health response that includes testing, for example efforts to control the COVID-19 pandemic. We have focused on self-reporting and contact-tracing, but the challenges are generic when there are distinct, but potentially undistinguished, primary and secondary recruiting processes. For example, more active general-population surveillance could still qualify as the primary recruitment in our model, as long as it were random with respect to intervention status. Likewise, geographic follow-up could be a secondary process, as long as intervention status were correlated with the secondary process (*e*.*g*. for dengue, adjacent households followed up, as long as vector control reached some areas and not others).

This analysis of the conventional TND and hybrid design under outbreak conditions does not consider other possible sources of bias, such as different exposure risk between groups, testing errors, or errors in ascertaining vaccine status. Further investigation of the reliability of these studies for estimating effectiveness during outbreaks remains critical. However, as we have shown, use of this design in an outbreak setting will need to account for the realities of control activities and plan to collect data on testing route or otherwise accommodate the mix of recruitment routes.

Adoption of a TND or hybrid design to evaluate a new vaccine in DRC may increase pressure for similar studies that do not have an explicit, randomised control group in future outbreaks of similarly highly pathogenic diseases. Understanding the biases and limitations of these designs will therefore be critical to evaluate vaccines that are currently being developed against these pathogens.

## Data Availability

The results are simulation-based. All code for those simulations can be found in the repository.

https://gitlab.com/ebovac/tncc_math

## Acknowledgements

We thank Peter G. Smith and Nick Jewell for useful discussions during the development of this model and analysis.

This work was supported by the Innovative Medicines Initiative 2 (IMI2) Joint Undertaking between European Union Horizon 2020 Research and Innovation Programme and the European Federation of Pharmaceutical Industries and Associations [EBOVAC3: grant number 800176]; Department of Health and Social Care using UK Aid funding managed by the NIHR [VEEPED: PR-OD-1017-20007]; National Institutes of Health, National Institute of General Medical Sciences [U54 GM111274]; and HDR UK Innovation Fellowship [grant MR/S003975/1 to R.M.E.]. The views expressed in this publication are those of the authors and not necessarily those of the Department of Health and Social Care.

