## Supplemental Info for "Potential Test-Negative Design Study Bias in Outbreak Settings: Application to Ebola vaccination in Democratic Republic of Congo"

*correspondence to: carl.pearson {at} lshtm.ac.uk

22 December, 2020

**S8 Alternative Scenario Translation** 42

### S1 Overview

A Test-Negative Design (TND) study has been proposed to evaluate a new Ebola Virus Disease (EVD) vaccine during the then-ongoing epidemic in Eastern Democratic Republic of Congo (DRC). The main text discussed a model of such a study: we described a population of individuals i) who heterogeneously receive a study vaccine, ii) some of whom subsequently exhibit symptoms of EVD or have contact with a known EVD case, and thus iii) are identified by either self-reporting and contact-tracing processes as part of an outbreak response, and finally iv) are tested for EVD, making them recruitable for a TND study of that vaccine. Particularly, we explored the potential for bias due to route of recruitment into the study, and due to heterogeneity in vaccine distribution. As we noted in the discussion, TND studies could be used to evaluate other kinds interventions.

To support application of the model to other contexts, we use more general terms in this Supplement (Section S2). Using those terms, we provide derivations of equations quoted in the main text (Sections S3-S6), additional results (Section S7), and translate the model to another example (Section S8).

### S2 Generalisation of Nomenclature

We consider a generic *study intervention*. The study intervention is in addition to any other outbreak control measures that might be ongoing. As in the main text, this study intervention is heterogeneously distributed at an individual level, which we represent with targeted status and intervention coverage. The TND study goal is to estimate the intervention efficacy; this observational estimate is often called the *effectiveness*.

The main text discusses self-reporting and contact-tracing as particular recruiting routes for EVD, which more generally are a random *primary* process and a reactive *secondary* process, respectively; we also use these qualifiers to distinguish the associated exposure processes to the pathogen of interest. For both routes, we still assume a highly sensitive and specific test for identifying infections with the target pathogen. For EVD, recruitment is symptom-related, but it does not have to be for all pathogens, so here we discuss intervention efficacy in terms of infection rather than disease.

The key assumptions in this model are:

- exposure to the target pathogen is identical for all individuals
- if infected by exposure, the probability of detection by the primary process is identical for all individuals
- the rate of exposure to any other pathogens that could result in testing (and therefore recruitment) is the same for all individuals
- all secondary recruits associated with an initial primary case have the same targeted status as that primary case
- the secondary transmission and recruitment process is identical for all individuals

#### S2.1 Model Diagram

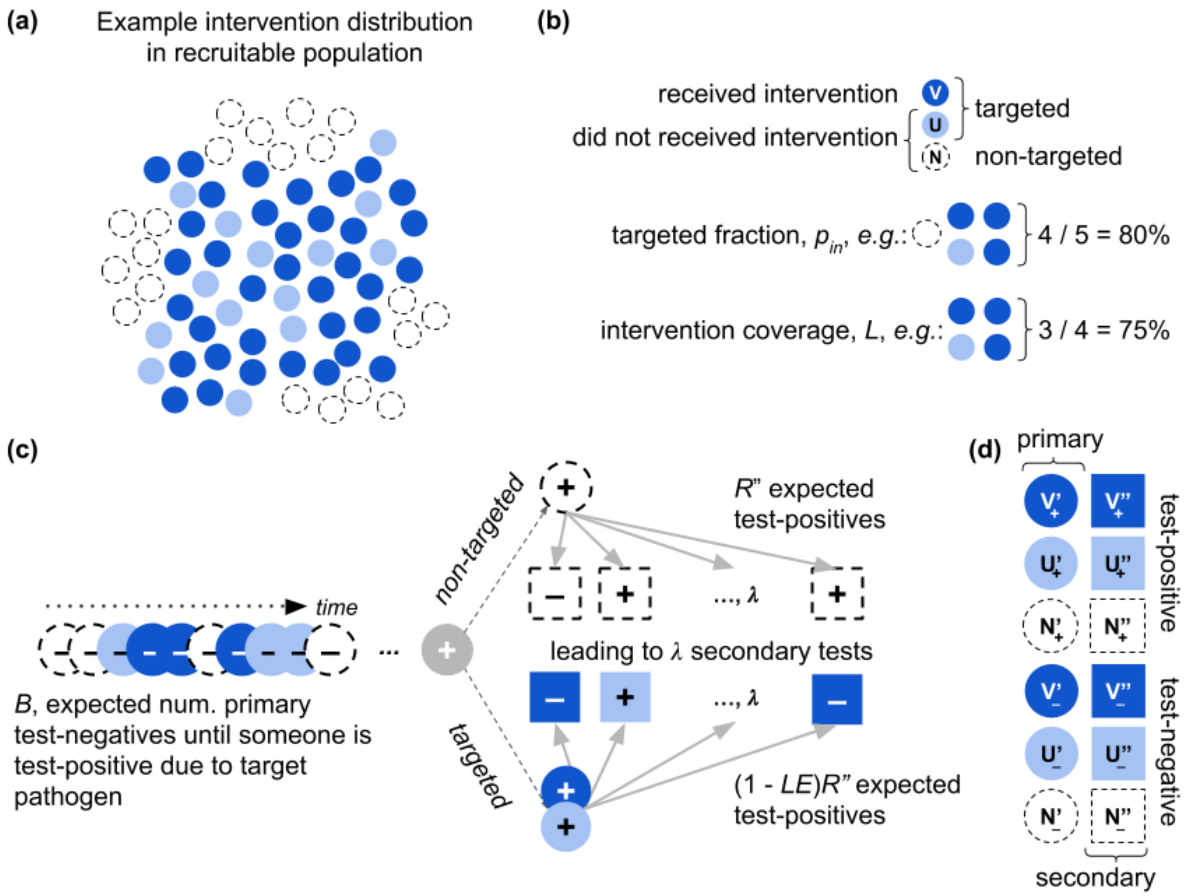

Figure S1: **Model Diagram**. This diagram illustrates the conceptual model of the population and recruitment process, showing: how the intervention might be distributed in the population (panel (a)), with the corresponding intervention term definitions for *targeted fraction*, $p_{\text{in}}$, and *intervention coverage*, $L$ (panel (b)); and how the primary process proceeds by finding an expected number of test-negatives, $B$, until finding a test-positive case, which initiates the secondary process leading to an expected number of additional tests, $\lambda$, of which $R''$ are expected to be test-positive in the absence of the study intervention (panel (c)), with the corresponding to summary categories of different routes to recruitment into the study (panel (d)).

#### S2.2 Population Heterogeneity

There are two groups within the recruitable population: *non-targeted* and *targeted*. Individuals in the non-targeted group do not receive the study intervention. Targeted individuals randomly receive the intervention with some probability. Aside from targeted and intervention status, individuals are identical: they have the same exposure risk for the target pathogen, same primary testing rate given exposure and non-exposure, and same secondary distribution and probability of exposing those individuals.

In the equations that follow, we label counts of individuals corresponding to their targeted and intervention status. The totals of these categories determine the population heterogeneity characteristics. Total numbers are:

$V$**,** the number of targeted individuals that received the intervention

$U$**,** the number of targeted individuals that did not receive the intervention

$C$**,** of the recruitable population, those that are targeted; $C = V + U$

$N$**,** individuals that were not targeted

$T$**,** total potentially recruitable study population; $T = C + N$

$p_{\mathbf{in}}$**,** the targeted fraction for the study, $p_{\text{in}} = \frac{C}{T}$; used as targeted probability for individuals

$L$**,** the intervention coverage in the targeted population, $L = \frac{V}{C}$; used as the probability targeted individuals have the intervention

#### S2.3 Exposure, Protection, Infection, and Recruitment

During the outbreak, individuals $\{V, U, N\}$ are potentially exposed and tested, and thus recruited in the study, via two routes: i) the primary route, where individuals are exposed and tested randomly, independent of any association with an identified case; ii) the secondary route, where individuals are exposed by and tested because of connection with an identified case.

Test-positive individuals found by the primary process and those they associate with as detect by the secondary process have the same targeted status in the model. Therefore primary test-positives from the targeted population only interact via the secondary process with other targeted individuals, and likewise for non-targeted primary test-positives.

Relative to transmission of the target pathogen, all infections discovered by secondary route are assumed to result from exposures due to the associated primary case. Primary cases are assumed to result from a random exposure process (*i.e.* exposing individuals of the different types according to the relative proportions in the population).

All exposures result in infections, unless protected by the study intervention; potential infections prevented by other outbreak response measures (which benefit all individuals in the study population equally) are assumed to not be exposures in the context of the model. For both primary and secondary exposures, an individual that has received the intervention may avoid infection with probability corresponding to the study intervention efficacy. Exposed individuals who have not received the intervention become infected.

The intervention is also assumed to have no impact on infections that are not by the target pathogen, even where those infections could lead to a test. Finally, neither process is assumed to observe all of the relevant events (*i.e.*, those that lead to primary or secondary recruitment) that occur, just that observation rates do not differ by targeted status.

#### S2.4 Recruitment-Related Parameters

These real-world infection and testing processes are stochastic, and we consider the asymptotic expected (*i.e.*, average) outcomes of these processes. We define the following set of parameters corresponding to expected counts of recruitment-related events per primary test-positive, and the probability of preventing infection.

**$B+1$:** the expected number of tests from the primary process required to find a single test-positive case; *i.e.*, on average, for $B+1$ individuals tested via primary route, $B$ are test-negative and 1 is test-positive.

**$\lambda$:** the expected number of tests from the secondary process, for each case identified by the primary process.

**$R''$:** the expected number of test-positives in the contacts of identified cases when there is no intervention.

**$E$:** the intervention efficacy; *i.e.*, the probability that an individual who has received the intervention will avoid infection when exposed (relative to an individual without the intervention) either via the primary or secondary routes

#### S2.5 Summary Recruitment Categories

In addition to the targeted and intervention states, we denote individuals by their routes to testing and test outcomes, with super- and subscripts respectively. These are summarized earlier in Fig. S1, and as follows:

**$T'_+$, $N'_+$, $C'_+=V'_++U'_+$:** among corresponding individuals: testing positive, recruited via primary process

**$T''_+$, $N''_+$, $C''_+=V''_++U''_+$:** from corresponding populations, testing positive, recruited via secondary process

**$T'_-$, $N'_-$, $C'_-=V'_-+U'_-$:** from corresponding populations, testing negative, recruited via primary process

**$T''_-$, $N''_-$, $C''_-=V''_-+U''_-$:** from corresponding populations, testing negative, recruited via secondary process

#### S2.6 Expanded TND Intervention Effectiveness Estimator

The conventional TND effectiveness estimator stratifies the recruited population into four groups, made from two binary distinctions: received the intervention versus not, and test-positive for the pathogen of interest versus test-negative. That effectiveness estimator is:

$$\begin{array}{c}\text{estimated}\\\text{effectiveness}\end{array} = 1 - \frac{\#\ \text{intervention, test-positive}}{\#\ \text{non-intervention, test-positive}} \times \frac{\#\ \text{non-intervention, test-negative}}{\#\ \text{intervention, test-negative}}$$

In terms of the types of individuals defined at the end of Section S2.3, this equation becomes:

$$\hat{E} = 1 - \frac{V'_+ + V''_+}{N'_+ + N''_+ + U'_+ + U''_+} \times \frac{N'_- + U'_- + N''_- + U''_-}{V'_- + V''_-} \tag{S1}$$

which can be thought of as two odds of having received the intervention, each conditional on the two possible test outcomes. We will refer to these as the test-positive odds and the test-negative odds.

### S3 Effectiveness Estimator Bias

To determine the potential bias of $\hat{E}$ for a TND study in an outbreak setting, we need to translate Eq. S1 from being in terms of total individual counts, into the expected totals, given the study target intervention and outbreak conditions. We will examine each of the two odds terms in turn, then combine the results.

#### S3.1 Test-Positive Odds

Starting with the test-positive odds term, we first factorise (by $C'_+$, or $N'_+$), such that most terms are expressed as proportions:

$$\frac{V'_+ + V''_+}{N'_+ + N''_+ + U'_+ + U''_+} = \frac{C'_+\left(\frac{V'_+}{C'_+} + \frac{V''_+}{C'_+}\right)}{N'_+\left(1 + \frac{N''_+}{N'_+}\right) + C'_+\left(\frac{U'_+}{C'_+} + \frac{U''_+}{C'_+}\right)} \tag{S2}$$

Recall we defined $R''$ as the expected number of cases that would be found via the secondary route among individuals that had not received the study intervention (*i.e.*, non-targeted individuals or individuals outside of the recruitable population). The proportion $\frac{N''_+}{N'_+}$ is total number of secondary cases over the total number of primary cases (among non-targeted individuals), which is also the average number of secondary cases per primary case, so we can substitute $R'' = \frac{N''_+}{N'_+}$. Since we have restricted the study to a scenario where only a single primary case exposes secondary cases, there is neither indirect protection or force of infection from multiple sources. Thus, amongst targeted individuals (*i.e.* those in $C$), $R''$ will be reduced on average by the probability that exposed individuals are intervention recipients and protected. This probability is equal to coverage, $L$, multiplied by the true efficacy, $E$. Therefore, $\frac{C''_+}{C'_+} = R''(1-LE)$ in the targeted population.

Using these substitutions and introducing some identity multipliers, $1 = \frac{C''_+}{C''_+}$, we can rewrite Eq. S2 as:

$$\frac{C'_+\left(\frac{V'_+}{C'_+} + \frac{C''_+}{C''_+}\frac{V''_+}{C'_+}\right)}{N'_+\left(1 + \frac{N''_+}{N'_+}\right) + C'_+\left(\frac{U'_+}{C'_+} + \frac{C''_+}{C''_+}\frac{U''_+}{C'_+}\right)} = \frac{C'_+\left(\frac{V'_+}{C'_+} + \frac{V''_+}{C''_+}R''(1-LE)\right)}{N'_+\left(1 + R''\right) + C'_+\left(\frac{U'_+}{C'_+} + \frac{U''_+}{C''_+}R''(1-LE)\right)} \tag{S3}$$

Next, we show how the proportions between the counts of the targeted individuals can be substituted to express the odds in terms of model parameters. Starting with $\frac{V'_+}{C'_+}$: this is the probability of an individual receiving the intervention, conditional on there being an initial infection in the targeted population. Though the exposure probabilities are the same between targeted and non-targeted populations, if $LE > 0$, the probability that an exposure results in an infection is lower. Given an exposure event:

$$\begin{aligned}\frac{V'_+}{C'_+} &= P(\text{received intervention} \mid \text{is infected \& targeted}) = P(i \in V|+, i \in C)\\ &= \frac{P(+|i \in V) \times P(i \in V|i \in C)}{P(+|i \in C)} = \frac{(1-E)\times L}{(1-L) + L(1-E)} = \frac{(1-E)L}{1-LE}\end{aligned}$$

We can use the same logic for the secondary exposures: conditional on a secondary exposure that could result in infection, the same relationship applies. To solve for $\frac{U'_+}{C'_+}$, we take the complements. Therefore, these four ratios are:

$$\begin{aligned} \frac{V'_+}{C'_+} = \frac{V''_+}{C''_+} &= \frac{(1-E)L}{1-LE} \\ \frac{U'_+}{C'_+} = \frac{U''_+}{C''_+} &= \frac{1-L}{1-LE} \end{aligned} \tag{S4}$$

which means we can further substitute in Eq. S3:

$$\begin{aligned} \frac{C'_+\left(\frac{V'_+}{C'_+} + \frac{V''_+}{C''_+}R''(1-LE)\right)}{N'_+(1+R'') + C'_+\left(\frac{U'_+}{C'_+} + \frac{U''_+}{C''_+}R''(1-LE)\right)} &= \frac{C'_+\frac{V'_+}{C'_+}(1+R''(1-LE))}{N'_+(1+R'') + C'_+\frac{U'_+}{C'_+}(1+R''(1-LE))} \\ &= \frac{C'_+\frac{(1-E)L}{1-LE}(1+R''(1-LE))}{N'_+(1+R'') + C'_+\frac{1-L}{1-LE}(1+R''(1-LE))} \\ &= \frac{\frac{C'_+}{T'_+}\frac{(1-E)L}{1-LE}}{\frac{N'_+}{T'_+}\frac{(1+R'')}{1+R''(1-LE)} + \frac{C'_+}{T'_+}\frac{1-L}{1-LE}} \end{aligned} \tag{S5}$$

Like for determining the fraction of infections in targeted individuals that did or did not receive the intervention (Eq. S3.1), we can also use Bayes Theorem to find the relative fractions of infections that occurred in individuals that were or were not targeted, $\frac{C'_+}{T'_+}$ and $\frac{N'_+}{T'_+}$:

$$\begin{aligned} \frac{C'_+}{T'_+} &= P(i \in C|+) = \frac{P(+|i \in C)P(i \in C)}{P(+)} = \frac{(1-LE)p_{\text{in}}}{(1-p_{\text{in}}) + (1-LE)p_{\text{in}}} &&= \frac{(1-LE)p_{\text{in}}}{1-LEp_{\text{in}}} \\ \frac{N'_+}{T'_+} &= P(i \in N|+) = 1 - P(i \in C|+) &&= \frac{1-p_{\text{in}}}{1-LEp_{\text{in}}} \end{aligned} \tag{S6}$$

which means that,

$$\begin{aligned} \frac{\frac{C'_+}{T'_+}\frac{(1-E)L}{1-LE}}{\frac{N'_+}{T'_+}\frac{(1+R'')}{1+R''(1-LE)} + \frac{C'_+}{T'_+}\frac{1-L}{1-LE}} &= \frac{1-LEp_{\text{in}}}{1-LEp_{\text{in}}} \frac{(1-LE)p_{\text{in}}\frac{(1-E)L}{1-LE}}{(1-p_{\text{in}})\frac{(1+R'')}{1+R''(1-LE)} + (1-LE)p_{\text{in}}\frac{1-L}{1-LE}} \\ &= \frac{p_{\text{in}}(1-E)L}{(1-p_{\text{in}})\frac{(1+R'')}{1+R''(1-LE)} + p_{\text{in}}(1-L)} \end{aligned} \tag{S7}$$

and therefore,

$$\frac{V'_+ + V_+}{N'_+ + N_+ + U'_+ + U_+} = \frac{p_{\text{in}}(1-E)L}{(1-p_{\text{in}})\frac{(1+R'')}{1+R''(1-LE)} + p_{\text{in}}(1-L)} \tag{S8}$$

In conventional TND studies there is no secondary recruitment. If secondary recruitment were eliminated under outbreak circumstances, that would imply that $\lambda \to 0$, which also means that $R'' \to 0$. During outbreaks, the secondary process would still occur as part of the response (*i.e.* there would both testing and case-finding), but the people identified would not be recruited. Under that limit:

$$
\lim_{R''\to 0} \frac{1+R''}{1+R''(1-LE)} = 1
$$

$$
\lim_{R''\to 0} \frac{Lp_{\text{in}}(1-E)}{(1-p_{\text{in}})\frac{1+R''}{1+R''(1-LE)} + p_{\text{in}} - Lp_{\text{in}}} = \frac{Lp_{\text{in}}(1-E)}{1-Lp_{\text{in}}} \tag{S9}
$$

which suggests a useful re-arrangement of the final form of the test-positive odds, so it has a clear separation of the terms which appear in the unbiased estimator (*i.e.* primary recruiting only) and the remaining factors:

$$
\begin{aligned}
\frac{Lp_{\text{in}}(1-E)}{(1-p_{\text{in}})\frac{1+R''}{1+R''(1-LE)} + p_{\text{in}}(1-L)} &= \frac{Lp_{\text{in}}(1-E)}{(1-p_{\text{in}})\frac{1+R''}{1+R''(1-LE)} + p_{\text{in}} - Lp_{\text{in}} + 1 - 1} \\
\text{obtain } 1 - Lp_{\text{in}} \text{ term to factor out...} &= \frac{Lp_{\text{in}}(1-E)}{(1-p_{\text{in}})\frac{1+R''}{1+R''(1-LE)} - (1-p_{\text{in}}) + (1-Lp_{\text{in}})} \\
\text{factor other term...} &= \frac{Lp_{\text{in}}(1-E)}{(1-Lp_{\text{in}}) + (1-p_{\text{in}})\left(\frac{1+R''}{1+R''(1-LE)} - 1\right)} \\
\text{simplify other term...} &= \frac{Lp_{\text{in}}(1-E)}{(1-Lp_{\text{in}}) + (1-p_{\text{in}})\left(\frac{LER''}{1+R''(1-LE)}\right)} \\
\text{factor out target terms...} &= \frac{Lp_{\text{in}}(1-E)}{1-Lp_{\text{in}}}\left[1 + \frac{ER''}{1+R''(1-LE)}\frac{L(1-p_{\text{in}})}{1-Lp_{\text{in}}}\right]^{-1}
\end{aligned} \tag{S10}
$$

In Eq. S10, we now have only terms that describe the intervention (targeted and coverage probabilities, $p_{\text{in}}$ and $L$, and efficacy $E$) and epidemiology ($R''$).

#### S3.2 Test-Negative Odds

We assume that the testing criteria for the secondary process is not affected by the presence of the intervention. For example, a contact-tracing-related criteria might be principally about high-risk interactions rather than particular symptoms, or the symptom threshold might be sufficiently relaxed that almost all contacts meet it. Similarly, a purely geographical criteria would be unaffected by presence or absence of the intervention. Thus, in our model all the prevented secondary infections (via true intervention efficacy $E$) are still recruited by the secondary process as test-negatives. This is a bounding assumption; see the end of this section for relaxing this assumption.

Turning to the test-negative odds, we first replace the primary test-negatives by the contribution from $B$, the average number of test-negatives per test-positive via the primary route. Given that definition, the total number of primary test-negatives is $T'_- = BT'_+$. Because the intervention has no effect on the causes that lead to testing negative via the primary route, the representation of individuals follows their proportions in the population:

$$
\frac{N'_- + U'_- + N''_- + U''_-}{V'_- + V''_-} = \frac{BT'_+(1-Lp_{\text{in}}) + N''_- + U''_-}{BT'_+Lp_{\text{in}} + V''_-} \tag{S11}
$$

As with the test-positives odds, we can factorise and introduce identity multiples to re-arrange into terms that we can then use Bayes Theorem to replace with model parameters:

$$\begin{aligned}\frac{BT'_+(1-Lp_{\text{in}}) + N''_- + U''_-}{BT'_+Lp_{\text{in}} + V''_-} &= \frac{B(1-Lp_{\text{in}}) + \frac{1}{T'_+}\left(N''_- + U''_-\right)}{BLp_{\text{in}} + \frac{V''_-}{T'_+}} \\ &= \frac{B(1-Lp_{\text{in}}) + \left(\frac{N'_+}{T'_+}\frac{N''_-}{N'_+} + \frac{C'_+}{T'_+}\frac{U''_-}{C'_+}\right)}{BLp_{\text{in}} + \frac{C'_+}{T'_+}\frac{V''_-}{C'_+}}\end{aligned} \tag{S12}$$

We can use the targeted and non-targeted fractions of primary test-positives, $\frac{C'_+}{T'_+}$ and $\frac{N'_+}{T'_+}$, from refactoring the test-positive odds (Eq. S6).

Amongst non-targeted individuals, on average $\lambda - R''$ of recruits from the secondary route will be test-negative. This means that $\frac{N''_-}{N'_+} = \lambda - R''$.

This definition also implies that the exposed proportion is $p_t = \frac{R''}{\lambda}$ because $R''$ individuals are infected per $\lambda$ secondary individuals. The complementary non-exposed proportion is therefore $1 - p_t = \frac{\lambda - R''}{\lambda}$. This value is like a transmission probability, though that interpretation should be used with caution: the denominator is determined by the secondary observation process, and thus the proportion may not clearly translate to the biological process probability.

Also by definition, amongst targeted individuals, only $1 - LE$ of the exposed individuals are infected, therefore:

$$\frac{C''_-}{C'_+} = (1 - p_t(1 - LE))\lambda = \lambda - R''(1 - LE)$$

We again use Bayes Theorem to translate these ratios into model parameter expressions.

$$\begin{aligned}\frac{U''_-}{C''_-} &= P(\text{is unvaccinated} \mid \text{is not infected \& targeted}) = P(i \in U|-, i \in C) = \frac{P(-|i \in U)P(i \in U|i \in C)}{P(-|i \in C)} \\ &= \frac{\frac{\lambda - R''}{\lambda}(1-L)}{\frac{\lambda - R''}{\lambda}(1-L) + L\left(\frac{\lambda - R''}{\lambda} + \frac{R''}{\lambda}E\right)} = \frac{(\lambda - R'')(1-L)}{\lambda - R''(1-LE)} \\ \frac{V''_-}{C''_-} &= 1 - P(i \in U|-, i \in C) = \frac{(\lambda - (1-E)R'')L}{\lambda - (1-LE)R''} \\ \frac{U''_-}{C'_+} &= \frac{U''_-}{C''_-}\frac{C''_-}{C'_+} = (\lambda - R'')(1-L) \\ \frac{V''_-}{C'_+} &= (\lambda - (1-E)R'')L\end{aligned} \tag{S13}$$

Substituting these into the for the appropriate ratios, we obtain:

$$\begin{aligned}\frac{B(1-Lp_{\text{in}}) + \left(\frac{N'_+}{T'_+}\frac{N_-}{N'_+} + \frac{C'_+}{T'_+}\frac{U_-}{C'_+}\right)}{BLp_{\text{in}} + \frac{C'_+}{T'_+}\frac{V_-}{C'_+}} &= \frac{B(1-Lp_{\text{in}}) + \left(\frac{1-p_{\text{in}}}{1-LEp_{\text{in}}}(\lambda - R'') + \frac{(1-LE)p_{\text{in}}}{1-LEp_{\text{in}}}(\lambda - R'')(1-L)\right)}{BLp_{\text{in}} + \frac{(1-LE)p_{\text{in}}}{1-LEp_{\text{in}}}(\lambda - (1-E)R'')L} \\ \frac{N'_- + U'_- + N_- + U_-}{V'_- + V_-} &= \frac{B(1-Lp_{\text{in}}) + \left(1 - Lp_{\text{in}}\frac{1-LE}{1-LEp_{\text{in}}}\right)(\lambda - R'')}{BLp_{\text{in}} + Lp_{\text{in}}\frac{1-LE}{1-LEp_{\text{in}}}(\lambda - R'' + ER'')}\end{aligned} \tag{S14}$$

As in Section S3.1, for the conventional TND assumptions, $\lambda \to 0$ and $R'' \to 0$. Again, it is not that the secondary process ceases, but just that recruitment via that route is disallowed. Enforcing those constraints:

$$\lim_{\lambda\to 0}\frac{B(1-Lp_{\text{in}})+\left(1-Lp_{\text{in}}\frac{1-LE}{1-LEp_{\text{in}}}\right)(\lambda-R'')}{BLp_{\text{in}}+Lp_{\text{in}}\frac{1-LE}{1-LEp_{\text{in}}}(\lambda-R''+ER'')}=\frac{B(1-Lp_{\text{in}})+\left(1-Lp_{\text{in}}\frac{1-LE}{1-LEp_{\text{in}}}\right)0}{BLp_{\text{in}}+Lp_{\text{in}}\frac{1-LE}{1-LEp_{\text{in}}}0}$$
$$=\frac{1-Lp_{\text{in}}}{Lp_{\text{in}}} \tag{S15}$$

As with test-positives odds, we can refactor in terms of the conventional TND limit:

$$\begin{aligned}\frac{B(1-Lp_{\text{in}})+\left(1-Lp_{\text{in}}\frac{1-LE}{1-LEp_{\text{in}}}\right)(\lambda-R'')}{BLp_{\text{in}}+Lp_{\text{in}}\frac{1-LE}{1-LEp_{\text{in}}}(\lambda-R''+ER'')} &= \frac{1-Lp_{\text{in}}}{Lp_{\text{in}}}\frac{B+\frac{1-Lp_{\text{in}}\frac{1-LE}{1-LEp_{\text{in}}}}{1-Lp_{\text{in}}}(\lambda-R'')}{B+\frac{1-LE}{1-LEp_{\text{in}}}(\lambda-R''+ER'')}\\ &= \frac{1-Lp_{\text{in}}}{Lp_{\text{in}}}\frac{B+\frac{1-LEp_{\text{in}}-Lp_{\text{in}}(1-LE)}{(1-Lp_{\text{in}})(1-LEp_{\text{in}})}(\lambda-R'')}{B+\frac{1-LE}{1-LEp_{\text{in}}}(\lambda-R''+ER'')}\\ &= \frac{1-Lp_{\text{in}}}{Lp_{\text{in}}}\frac{B+\frac{1-Lp_{\text{in}}-LEp_{\text{in}}+L^2Ep_{\text{in}}}{1-Lp_{\text{in}}-LEp_{\text{in}}+L^2Ep_{\text{in}}^2}(\lambda-R'')}{B+\frac{1-LE}{1-LEp_{\text{in}}}(\lambda-R''+ER'')}\end{aligned} \tag{S16}$$

In Eq. S16, we now have only terms that describe the intervention ($p_{\text{in}}$, $L$, and $E$) and epidemiology ($R''$, $\lambda$, and $B$). Note that this term includes more of the model parameters than the test-positive odds (Eq. S10).

#### S3.3 Relaxing Assumption that Prevented Infections Remain Secondary Recruits

Earlier, we assumed that the secondary recruitment process was unperturbed by the study intervention. For a secondary process that is, for example, purely geographical because it concerns a pathogen that is highly asymptomatic (*e.g.*, neighbor-household testing for dengue), this assumption is consistent. Where it becomes less clearly acceptable as a simplification, is if there remains some disease- or symptom-based component to secondary recruitment.

In the main text, we focus on a vaccine study for EVD, where the primary process was self-reporting with multiple EVD-like symptoms leading to testing. The secondary process is nominally contact-tracing combined with a fever. We assumed that subjective fever would almost always be present for the contacts that avoided EVD infection because of the vaccine. In reality there would be some attack rate less than 100%.

If we define the proportion of people meeting a symptom-based component of the secondary process as $\alpha$ and ignore the zero-bias term (Eq. S15) as a coefficient, then Eq. S16 becomes:

$$\text{test-negative odds} \propto \frac{B+\frac{1-Lp_{\text{in}}-LEp_{\text{in}}+L^2Ep_{\text{in}}}{1-Lp_{\text{in}}-LEp_{\text{in}}+L^2Ep_{\text{in}}^2}(\lambda-R'')}{B+\frac{1-LE}{1-LEp_{\text{in}}}(\lambda-R''+E\alpha R'')} \tag{S17}$$

That is, of all the potential secondary recruits that could be added to test-negatives due to prevention of infection, only some exhibit the additional criteria. Note that there is no impact of relaxing this assumption on the test-positive odds.

If $\alpha \to 1$, *i.e.* everyone meets this extra criteria, we get Eq. S16. As $\alpha \to 0$, the denominator decreases, increasing the test-negative odds overall, and in turn reducing the estimated effectiveness. When $\alpha = 0$:

$$\text{test-negative odds} \propto \frac{B + \frac{1-Lp_{\text{in}}-LEp_{\text{in}}+L^2Ep_{\text{in}}}{1-Lp_{\text{in}}-LEp_{\text{in}}+L^2Ep_{\text{in}}^2}(\lambda - R'')}{B + \frac{1-LE}{1-LEp_{\text{in}}}(\lambda - R'')} \tag{S18}$$

In the case of $p_{\text{in}} = 1$, this reduces to unity. More generally,

$$\begin{aligned}
Lp_{\text{in}} \le L &\implies 1 - Lp_{\text{in}} \ge 1 - L \implies \frac{1-L}{1-Lp_{\text{in}}} \le 1 \\
&\implies 1 \ge p_{\text{in}}\frac{1-L}{1-Lp_{\text{in}}} \implies LE \ge LEp_{\text{in}}\frac{1-L}{1-Lp_{\text{in}}} \\
&\implies 1 - LE \le 1 - LEp_{\text{in}}\frac{1-L}{1-Lp_{\text{in}}} \implies 1 - LE \le \frac{1 - Lp_{\text{in}} - LEp_{\text{in}} - L^2Ep_{\text{in}}}{1 - Lp_{\text{in}}} \\
&\implies \frac{1-LE}{1-LEp_{\text{in}}} \le \frac{1 - Lp_{\text{in}} - LEp_{\text{in}} - L^2Ep_{\text{in}}}{(1 - LEp_{\text{in}})(1 - Lp_{\text{in}})} \\
&\implies B + \frac{1-LE}{1-LEp_{\text{in}}}(\lambda - R'') \le B + \frac{1 - Lp_{\text{in}} - LEp_{\text{in}} - L^2Ep_{\text{in}}}{(1 - LEp_{\text{in}})(1 - Lp_{\text{in}})}(\lambda - R'') \\
&\implies 1 \le \frac{B + \frac{1-Lp_{\text{in}}-LEp_{\text{in}}-L^2Ep_{\text{in}}}{(1-LEp_{\text{in}})(1-Lp_{\text{in}})}(\lambda - R'')}{B + \frac{1-LE}{1-LEp_{\text{in}}}(\lambda - R'')}
\end{aligned} \tag{S19}$$

This means that without any contribution from $\alpha$, test-negative odds biases increasingly towards underestimation as targeted fraction decreases. As $\alpha \to 1$, this effect is counteracted, but can in turn lead to overestimation of effectiveness.

### S4 Total Estimator Bias & Limiting Scenarios

Combining the test-positives odds and test-negatives odds:

$$\begin{aligned}
\hat{E} &= 1 - \frac{V'_+ + V''_+}{N'_+ + N''_+ + U'_+ + U''_+}\frac{N'_- + U'_- + N''_- + U''_-}{V'_- + V''_-} \\
&= 1 - \frac{Lp_{\text{in}}(1-E)}{1-Lp_{\text{in}}}\left[1 + \frac{ER''}{1 + R''(1-LE)}\frac{L(1-p_{\text{in}})}{1-Lp_{\text{in}}}\right]^{-1}\frac{1-Lp_{\text{in}}}{Lp_{\text{in}}}\frac{B + \frac{1-Lp_{\text{in}}-LEp_{\text{in}}+L^2Ep_{\text{in}}}{1-Lp_{\text{in}}-LEp_{\text{in}}+L^2Ep_{\text{in}}^2}(\lambda - R'')}{B + \frac{1-LE}{1-LEp_{\text{in}}}(\lambda - R'' + ER'')} \\
&= 1 - (1-E)\left[1 + \frac{ER''}{1 + R''(1-LE)}\frac{L(1-p_{\text{in}})}{1-Lp_{\text{in}}}\right]^{-1}\frac{B + \frac{1-Lp_{\text{in}}-LEp_{\text{in}}+L^2Ep_{\text{in}}}{1-Lp_{\text{in}}-LEp_{\text{in}}+L^2Ep_{\text{in}}^2}(\lambda - R'')}{B + \frac{1-LE}{1-LEp_{\text{in}}}(\lambda - R'' + ER'')}
\end{aligned} \tag{S20}$$

This suggests a different factorization:

$$\hat{E} = 1 - (1-E)\left[1 + \frac{E\frac{R''}{\lambda}\frac{\lambda}{B+1}(B+1)}{1 + \frac{R''}{\lambda}\frac{\lambda}{B+1}(B+1)(1-LE)}\frac{L(1-p_{\text{in}})}{1-Lp_{\text{in}}}\right]^{-1}\frac{1 + \frac{1-Lp_{\text{in}}-LEp_{\text{in}}+L^2Ep_{\text{in}}}{1-Lp_{\text{in}}-LEp_{\text{in}}+L^2Ep_{\text{in}}^2}\frac{\lambda}{B+1}\frac{B+1}{B}\left(1 - \frac{R''}{\lambda}\right)}{1 + \frac{1-LE}{1-LEp_{\text{in}}}\frac{\lambda}{B+1}\frac{B+1}{B}(1 - \frac{R''}{\lambda} + E\frac{R''}{\lambda})} \tag{S21}$$

We can define secondary test-positive fraction, $p_t = \frac{R''}{\lambda}$, the negative proportion of primary alerts, $f_- = \frac{B}{B+1}$, and relative rate of secondary recruitment to primary recruitment, $\rho = \frac{\lambda}{B+1}$. Noting that $B + 1 = (1 - f_-)^{-1}$ This yields:

$$
\hat{E} = 1 - (1-E)\left[1 + \frac{E\frac{p_t\rho}{1-f_-}}{1+\frac{p_t\rho}{1-f_-}(1-LE)}\frac{L(1-p_{\mathrm{in}})}{1-Lp_{\mathrm{in}}}\right]^{-1} \frac{1+\frac{1-Lp_{\mathrm{in}}-LEp_{\mathrm{in}}+L^2Ep_{\mathrm{in}}}{1-Lp_{\mathrm{in}}-LEp_{\mathrm{in}}+L^2Ep_{\mathrm{in}}^2}\frac{\rho}{f_-}(1-p_t)}{1+\frac{1-LE}{1-LEp_{\mathrm{in}}}\frac{\rho}{f_-}(1-p_t(1-E))} \tag{S22}
$$

We use this framing for all the main text results. This formulation highlights the important relative values within the model, while still maintaining terms that can be reasoned about and potentially measured. In this framing $\hat{E}$ is still a function of six variables, *i.e.* $\{E, L, p_{\mathrm{in}}, R'', \lambda, B\}$ *versus* $\{E, L, p_{\mathrm{in}}, p_t, \rho, f_-\}$.

In the following subsections, we show limiting conditions for the estimator with respect to the assorted parameters.

#### S4.1 True efficacy, $E$, limits

As the intervention tends toward either doing nothing ($E \to 0$) or perfect protection ($E \to 1$), the estimator bias tends to vanish.

$$
\begin{aligned}
\lim_{E\to 0}\hat{E} &= 1 - [1]^{-1}\frac{1+\frac{1-Lp_{\mathrm{in}}}{1-Lp_{\mathrm{in}}}\frac{\rho}{f_-}(1-p_t)}{1+\frac{\rho}{f_-}(1-p_t)} = 1 - 1 = 0 \\
\lim_{E\to 1}\hat{E} &= 1 - 0\,(\cdots) = 1
\end{aligned} \tag{S23}
$$

#### S4.2 Targeted fraction, $p_{\mathrm{in}}$, limits

$$
\begin{aligned}
\lim_{p_{\mathrm{in}}\to 0}\hat{E} &= 1 - (1-E)\left[1 + \frac{LE\frac{p_t\rho}{1-f_-}}{1+\frac{p_t\rho}{1-f_-}(1-LE)}\right]^{-1}\frac{1+\frac{1}{1}\frac{\rho}{f_-}(1-p_t)}{1+\frac{1-LE}{1}\frac{\rho}{f_-}(1-p_t(1-E))} \\
&= 1 - (1-E)\left[\frac{1+\frac{p_t\rho}{1-f_-}(1-LE)}{1+\frac{p_t\rho}{1-f_-}(1-LE)} + \frac{LE\frac{p_t\rho}{1-f_-}}{1+\frac{p_t\rho}{1-f_-}(1-LE)}\right]^{-1}\frac{1+\frac{\rho}{f_-}(1-p_t)}{1+(1-LE)\frac{\rho}{f_-}(1-p_t(1-E))} \\
&= 1 - (1-E)\left[\frac{1+\frac{p_t\rho}{1-f_-}}{1+\frac{p_t\rho}{1-f_-}(1-LE)}\right]^{-1}\frac{1+\frac{\rho}{f_-}(1-p_t)}{1+(1-LE)\frac{\rho}{f_-}(1-p_t(1-E))} \\
&= 1 - (1-E)\left[\frac{1+\frac{p_t\rho}{1-f_-}}{1+\frac{p_t\rho}{1-f_-}(1-LE)}\right]^{-1}\frac{1+\frac{\rho}{f_-}(1-p_t)}{1+(1-LE)\frac{\rho}{f_-}(1-p_t(1-E))} \\
&= 1 - (1-E)\frac{1+\frac{p_t\rho}{1-f_-}(1-LE)}{1+\frac{p_t\rho}{1-f_-}}\frac{1+\frac{\rho}{f_-}(1-p_t)}{1+(1-LE)\frac{\rho}{f_-}(1-p_t(1-E))}
\end{aligned} \tag{S24}
$$

$$
\begin{aligned}
\lim_{p_{\text{in}}\to 1} \hat{E} &= 1 - (1-E)\,[1]^{-1}\,\frac{B + \left[1 + \frac{L}{1-L}0\right](\lambda - R'')}{B + (\lambda - R'' + ER'')} \\
&= 1 - (1-E)\left[1 + 0\frac{E\frac{p_t\rho}{1-f_-}}{1 + \frac{p_t\rho}{1-f_-}(1-LE)}\right]^{-1}\frac{1 + \frac{1-L-LE+L^2E}{1-L-LE+L^2E}\frac{\rho}{f_-}(1-p_t)}{1 + \frac{1-LE}{1-LE}\frac{\rho}{f_-}(1-p_t(1-E))} \\
&= 1 - (1-E)\frac{1 + \frac{\rho}{f_-}(1-p_t)}{1 + \frac{\rho}{f_-}(1-p_t(1-E))} \\
&= 1 - (1-E)\frac{B + \lambda - R''}{B + \lambda - R'' + ER''} \\
&= 1 - (1-E)\frac{1 - \frac{R''}{B+\lambda}}{1 - \frac{R''}{B+\lambda}(1-E)}
\end{aligned} \tag{S25}
$$

The final factorization in Eq. S25 shows that, in the limit of perfect alignment of targeting and recruitment, the intervention coverage, $L$, is removed, and the bias depends only on the true efficacy, $E$, and a combination of epidemiological parameters: $\frac{R''}{B+\lambda}$. This relationship can be inverted; which allows us to determine the true efficacy in terms of the estimator value and other parameters:

$$
\begin{aligned}
1 - \hat{E} &= (1-E)\frac{1 - \frac{R''}{B+\lambda}}{1 - \frac{R''}{B+\lambda}(1-E)} \\
\left(1-\hat{E}\right)\left(1 - \frac{R''}{B+\lambda}(1-E)\right) &= (1-E)\left(1 - \frac{R''}{B+\lambda}\right) \\
\left(1-\hat{E}\right) &= (1-E)\left[1 - \frac{R''}{B+\lambda} + \left(1-\hat{E}\right)\frac{R''}{B+\lambda}\right] \\
(1-E) &= \left(1-\hat{E}\right)\left[1 - \hat{E}\frac{R''}{B+\lambda}\right]^{-1} \\
E &= 1 - \left(1-\hat{E}\right)\left[1 - \hat{E}\frac{R''}{B+\lambda}\right]^{-1} \\
E - \hat{E} &= (1-\hat{E})\left(1 - \left[1 - \hat{E}\frac{R''}{B+\lambda}\right]^{-1}\right) \\
E - \hat{E} &= -(1-\hat{E})\left(\frac{\hat{E}\frac{R''}{B+\lambda}}{1 - \hat{E}\frac{R''}{B+\lambda}}\right)
\end{aligned} \tag{S26}
$$

$\frac{R''}{B+\lambda}$ corresponds to a potentially measurable quantity; recall that in the model, $R''$ is the expected number of additional test-positives that are identified via the secondary process in a group without the intervention, and $B+\lambda$ is all the other tests (primary negatives and all secondary tests) per primary test-positive. Thus, $\frac{R''}{B+\lambda}$ is the fraction of secondary test-positives out of all non-index case-finding tests, when measuring in a non-intervention group. This value could be estimated from a comparable population without the intervention, or an upper limit could be estimated from data within the study population itself: the expected number of secondary cases in the intervention population is $R''(1-LE)$, so the measured non-primary test-positive fraction would be reduced maximally by a factor $(1-L)$ when the efficacy is perfect. Note that because this factor only has $B + \lambda$, we do not need to distinguish primary versus secondary test-negatives. If a test-negative from $\lambda$ was mistakenly assigned to $B$ (or *vice versa*), that would not change this factor.

Thus, if a study were able to achieve $p_{\text{in}} \approx 1$, it only need to be able to distinguish between primary and secondary test-positives to correctly bound the estimator.

##### S4.3 Secondary Case Recruitment, $p_t$, limits

If we consider secondary *case* limits, that is what proportion of secondary recruiting is test-positive, then we obtain:

$$\begin{aligned}
\lim_{p_t \to 0} \hat{E} &= 1 - (1-E)\left[1 + \frac{0}{1+0}\frac{L(1-p_{\text{in}})}{1-Lp_{\text{in}}}\right]^{-1} \frac{1 + \frac{1-Lp_{\text{in}}-LEp_{\text{in}}+L^2Ep_{\text{in}}}{1-Lp_{\text{in}}-LEp_{\text{in}}+L^2Ep_{\text{in}}^2}\frac{\rho}{f_-}}{1+\frac{1-LE}{1-LEp_{\text{in}}}\frac{\rho}{f_-}} \\
&= 1 - (1-E)\frac{1 + \frac{1-Lp_{\text{in}}-LEp_{\text{in}}+L^2Ep_{\text{in}}}{1-Lp_{\text{in}}-LEp_{\text{in}}+L^2Ep_{\text{in}}^2}\frac{\rho}{f_-}}{1+\frac{1-LE}{1-LEp_{\text{in}}}\frac{\rho}{f_-}} \\
\lim_{p_t \to 1} \hat{E} &= 1 - (1-E)\left[1 + \frac{E\frac{\rho}{1-f_-}}{1+\frac{\rho}{1-f_-}(1-LE)}\frac{L(1-p_{\text{in}})}{1-Lp_{\text{in}}}\right]^{-1} \frac{1 + \frac{1-Lp_{\text{in}}-LEp_{\text{in}}+L^2Ep_{\text{in}}}{1-Lp_{\text{in}}-LEp_{\text{in}}+L^2Ep_{\text{in}}^2}\frac{\rho}{f_-}0}{1+\frac{1-LE}{1-LEp_{\text{in}}}\frac{\rho}{f_-}E} \\
&= 1 - (1-E)\left[1 + \frac{E\frac{\rho}{1-f_-}}{1+\frac{\rho}{1-f_-}(1-LE)}\frac{L(1-p_{\text{in}})}{1-Lp_{\text{in}}}\right]^{-1} \frac{1}{1+\frac{1-LE}{1-LEp_{\text{in}}}\frac{\rho}{f_-}E}
\end{aligned} \tag{S27}$$

One way to interpret $p_t \to 1$ is that transmission probability (conditional on high risk contact) is going up. Another way to think about it is $\lambda$ coming down to meet $R''$; *i.e.*, the decision about whether to test a contact or not becoming more accurately linked to whether they were infected.

##### S4.4 Lower Limit on Secondary Relative Recruiting, $\rho$

As primary recruiting increasingly outweighs secondary recruiting (including both increasing primary recruitment and disallowing secondary recruitment), $\rho \to 0$. In this limit:

$$\begin{aligned}
\lim_{\rho \to 0} \hat{E} &= 1 - (1-E)\left[1 + \frac{0}{1+0}\frac{L(1-p_{\text{in}})}{1-Lp_{\text{in}}}\right]^{-1} \frac{1 + \frac{1-Lp_{\text{in}}-LEp_{\text{in}}+L^2Ep_{\text{in}}}{1-Lp_{\text{in}}-LEp_{\text{in}}+L^2Ep_{\text{in}}^2}\frac{0}{f_-}(1-p_t)}{1+\frac{1-LE}{1-LEp_{\text{in}}}\frac{0}{f_-}(1-p_t(1-E))} \\
&= 1 - (1-E)[1]^{-1}\frac{1}{1} = E
\end{aligned} \tag{S28}$$

Thus, for sufficiently high rate of primary recruitment leading to test-negatives, the bias goes to 0.

#### S5 Hybrid Study Design

Given that the bias in conventional design arises from aggregating the primary and secondary recruitment routes, we might expect that treating the recruitment routes as separate could limit this bias. If we consider the secondary recruitment population as a cohort study, then the conventional cohort design estimator is:

$$\begin{array}{c}\text{estimated}\\ \text{effectiveness}\end{array} = 1 - \frac{\text{attack rate in individuals receiving study intervention}}{\text{attack rate in remaining individuals}}$$

$$\hat{E} = 1 - \frac{V''_+}{V''} \times \frac{N''+U''}{N''_+ + U''_+} = 1 - \frac{V''_+}{N''_+ + U''_+} \times \frac{N''+U''}{V''} \tag{S29}$$

We make a simpler argument in the main text, but we can also use previously identified relationships to show this is unbiased. Recall Eq. S4 and additional definitions:

$$
\begin{aligned}
\frac{V'_+}{C'_+} = \frac{V''_+}{C''_+} &= \frac{(1-E)L}{1-LE} \\
\frac{U'_+}{C'_+} = \frac{U''_+}{C''_+} &= \frac{1-L}{1-LE} \\
\frac{N''}{N'_+} &= \lambda \\
\frac{V''}{C'_+} &= L\lambda \\
\frac{U''}{C'_+} &= (1-L)\lambda
\end{aligned}
\tag{S30}
$$

Using a similar approach as that for the TND estimator, and re-using those ratios:

$$
\begin{aligned}
\hat{E} &= 1 - \frac{\frac{C'_+}{T'_+}\frac{V''_+}{C'_+}}{\frac{N'_+}{T'_+}\frac{N''_+}{N'_+} + \frac{C'_+}{T'_+}\frac{U''_+}{C'_+}} \times \frac{\frac{N'_+}{T'_+}\frac{N''}{N'_+} + \frac{C'_+}{T'_+}\frac{U''}{C'_+}}{\frac{C'_+}{T'_+}\frac{V''}{C'_+}} \\
&= 1 - \frac{\frac{C''_+}{C'_+}\frac{V''_+}{C''_+}}{\frac{N'_+}{T'_+}\frac{N''_+}{N'_+} + \frac{C'_+}{T'_+}\frac{C''_+}{C'_+}\frac{U''_+}{C''_+}} \times \frac{\frac{N'_+}{T'_+}\lambda + \frac{C'_+}{T'_+}(1-L)\lambda}{L\lambda} \\
&= 1 - \frac{R''(1-LE)\frac{V'_+}{C'_+}}{\frac{N'_+}{T'_+}R'' + \frac{C'_+}{T'_+}R''(1-LE)\frac{U'_+}{C'_+}} \times \frac{\frac{N'_+}{T'_+} + \frac{C'_+}{T'_+}(1-L)}{L} \\
&= 1 - \frac{(1-E)L}{\frac{N'_+}{T'_+} + \frac{C'_+}{T'_+}(1-L)} \times \frac{\frac{N'_+}{T'_+} + \frac{C'_+}{T'_+}(1-L)}{L} \\
&= 1 - (1-E) = E
\end{aligned}
\tag{S31}
$$

Eq. S31 implies that if it were possible to observe secondary cases (out of secondary contacts) as a cohort, there would be no bias in such a study, regardless of heterogeneity in vaccine uptake. Another advantage of this study design is that there's no uncertainty about testing criteria (high risk contacts, regardless of symptoms), unlike the test-negative design (where symptoms may play a role in secondary recruitment).

#### S5.1 Hybrid Estimator

We can now consider combining the TND estimator and the cohort estimator:

$$
\hat{E} = 1 - \frac{\omega_{\mathrm{OR}}\mathrm{OR}_{\mathrm{TND}} + \omega_{\mathrm{RR}}\mathrm{RR}_{\mathrm{CS}}}{\sum \omega} \tag{S32}
$$

In this combination, the limiting condition for the TND of no secondary testing applies (all those individuals go into the cohort term) and thus the TND estimator is unbiased (per Eq. S28). We have just shown that the cohort study using only secondary recruitment is also unbiased (under our other assumptions). Thus, any weighted average of the terms, like Eq. S32, is also unbiased. Therefore, selection of these weights can optimize for other study features, such as power.

### S6 Translation of Limits to Recruitment Constraints for Conventional TND

#### S6.1 Attempting to Limit Recruitment to Targeted Population Only

Figures S2-S10 show the general response of the estimator to varying factors in the model. Each plot shows $p_{\text{in}} \in \{0.01, 0.1, 0.25, 0.5, 0.75, 0.9, 1\}$ (columns) and $p_t \in \{0.01, 0.1, 0.25, 0.5, 0.75, 0.9, 1\}$ (rows). Each plot shows one of the combinations of $\rho \in \{1/9, 1/3, 1\}$ and $f_- \in \{0.5, 0.75, 0.9\}$; $\rho$ is indicated at the top of each plot, $f_-$ on the right side.

These plots show some trends under specific conditions. In general, increasing targeted fraction decreases bias range, though not absolutely (*e.g.*, $\rho = 1/9, p_t = 1/4, f_- = 0.75$). Increasing coverage can shift bias towards underestimation or overestimation, depending $p_t$.

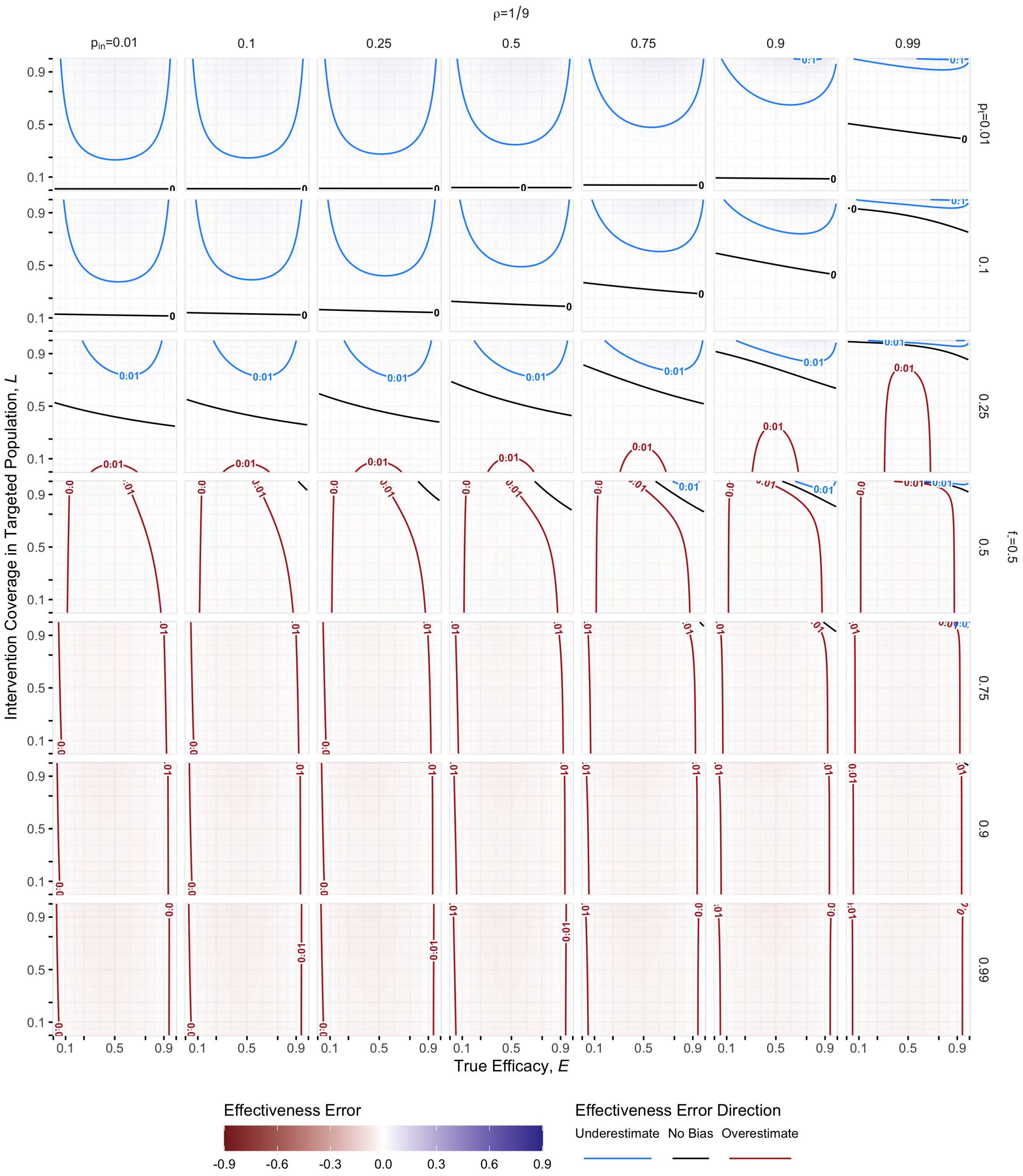

Figure S2: **General Bias Sensitivity, 1 of 9**: These series of plots show general sensitivity of the TND estimator to all of the model parameters. For each plot, the rightmost column corresponds to very high (99%) targeted fraction, which indicates the minimal bias surface when the study manages to maximize targeted fraction. Recall, $\rho = \frac{\lambda}{B+1}$ is the expected ratio of secondary to primary recuits; $p_t = R''/\lambda$ is the expected fraction of secondary recruits that test positive when no intervention is present; and $f_- = \frac{B}{B+1}$ is the expect fraction of primary recruits that are test-negative. *In this panel,* $\rho = 1/9$ *and* $f_- = 0.5$.

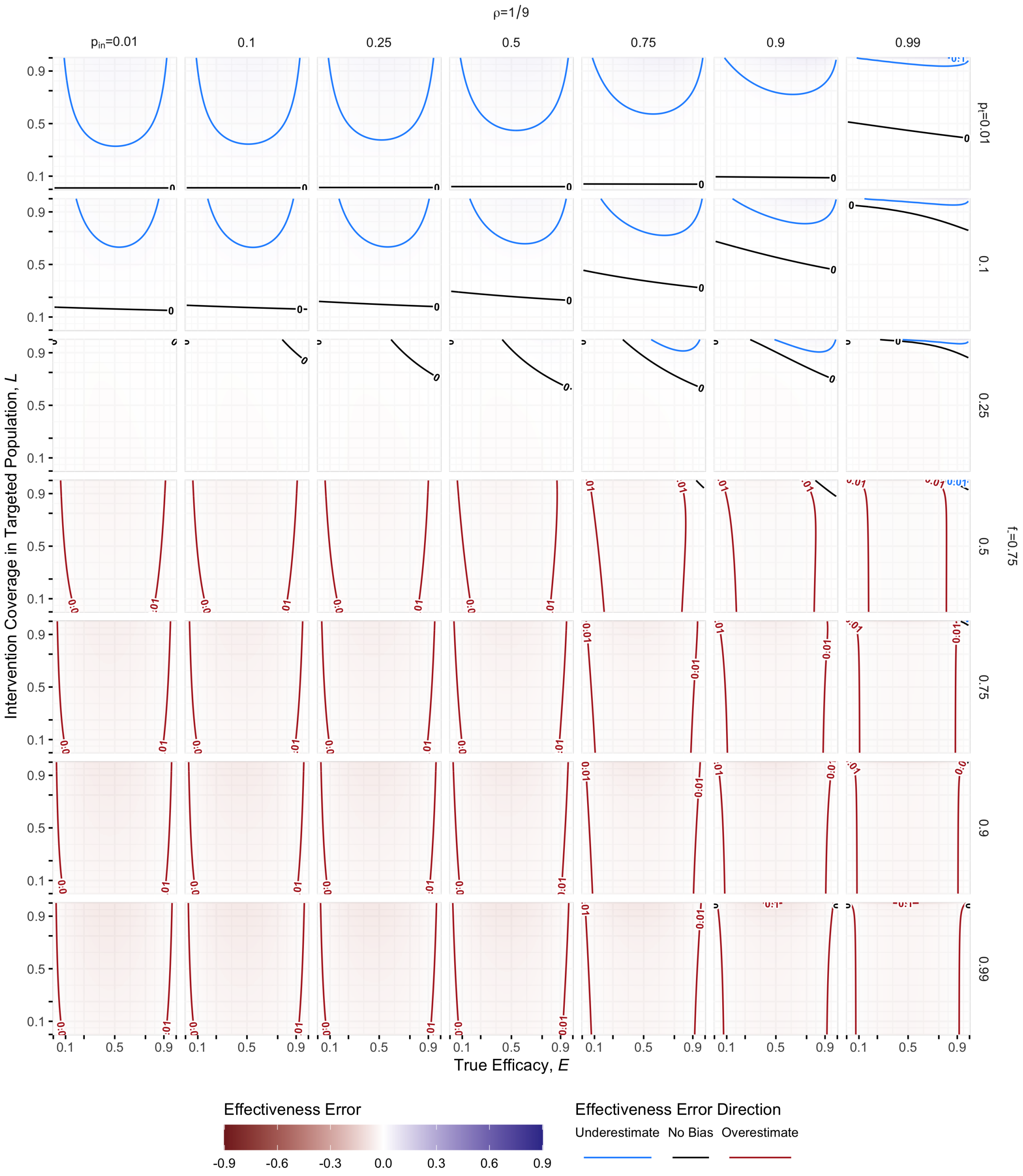

Figure S3: **General Bias Sensitivity. 2 of 9**: Recall, $\rho = \frac{\lambda}{B+1}$ is the expected ratio of secondary to primary recuits; $p_t = R''/\lambda$ is the expected fraction of secondary recruits that test positive when no intervention is present; and $f_- = \frac{B}{B+1}$ is the expect fraction of primary recruits that are test-negative. *In this panel,* $\rho = 1/9$ *and* $f_- = 0.75$.

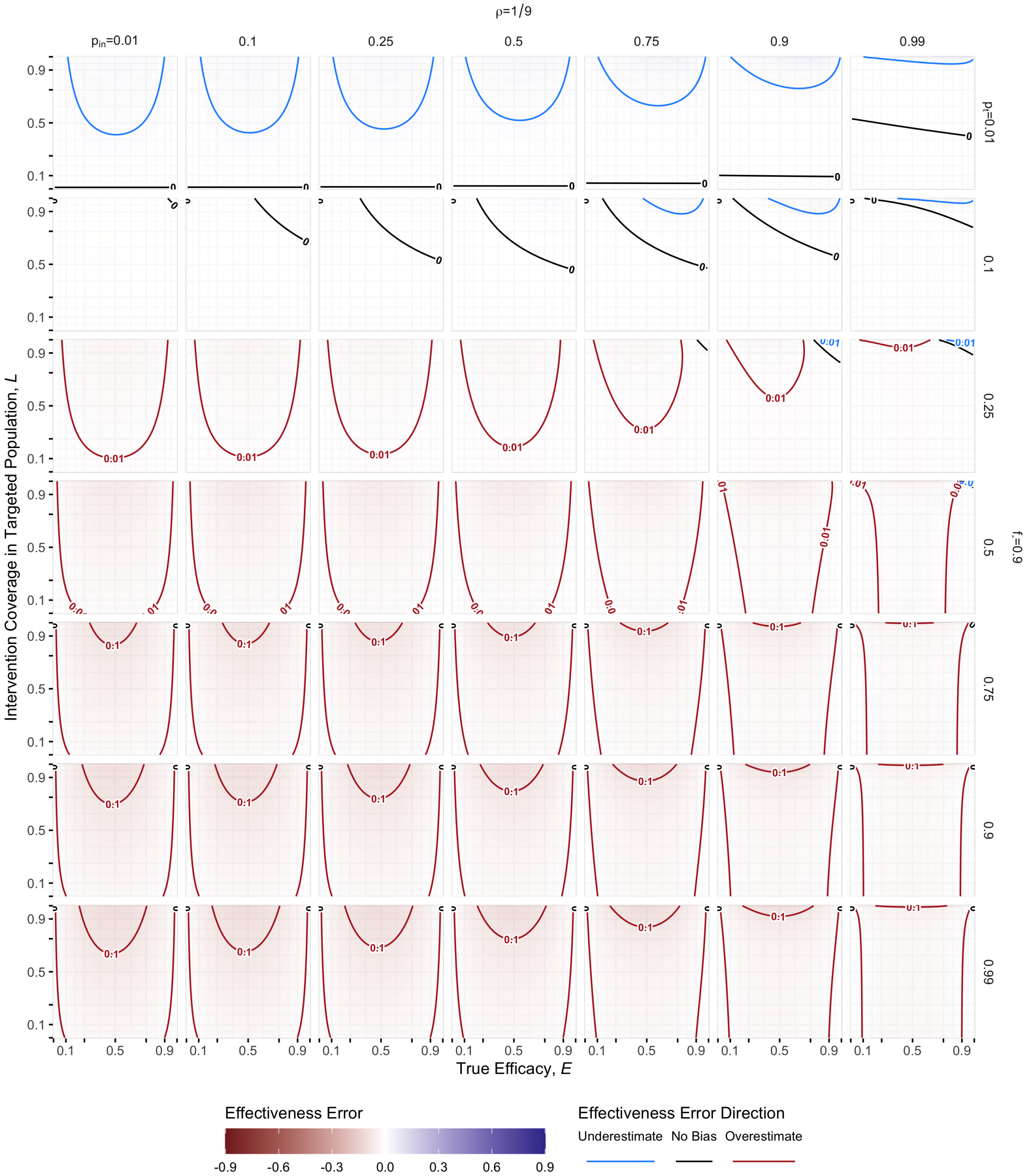

Figure S4: **General Bias Sensitivity. 3 of 9**: Recall, $\rho = \frac{\lambda}{B+1}$ is the expected ratio of secondary to primary recuits; $p_t = R''/\lambda$ is the expected fraction of secondary recruits that test positive when no intervention is present; and $f_- = \frac{B}{B+1}$ is the expect fraction of primary recruits that are test-negative. *In this panel,* $\rho = 1/9$ *and* $f_- = 0.9$.

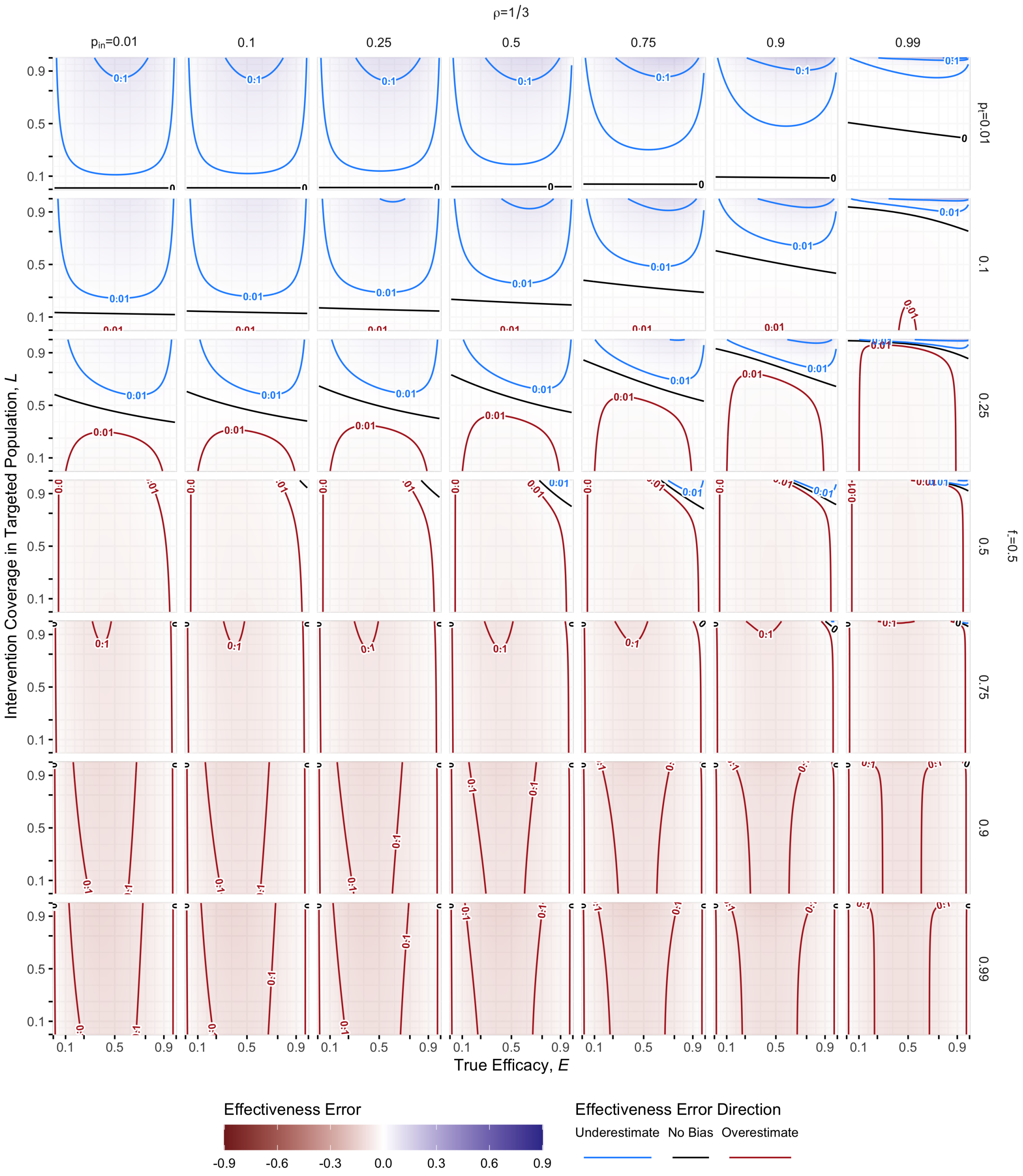

Figure S5: **General Bias Sensitivity. 4 of 9**: Recall, $\rho = \frac{\lambda}{B+1}$ is the expected ratio of secondary to primary recuits; $p_t = R''/\lambda$ is the expected fraction of secondary recruits that test positive when no intervention is present; and $f_- = \frac{B}{B+1}$ is the expect fraction of primary recruits that are test-negative. *In this panel,* $\rho = 1/3$ *and* $f_- = 0.5$.

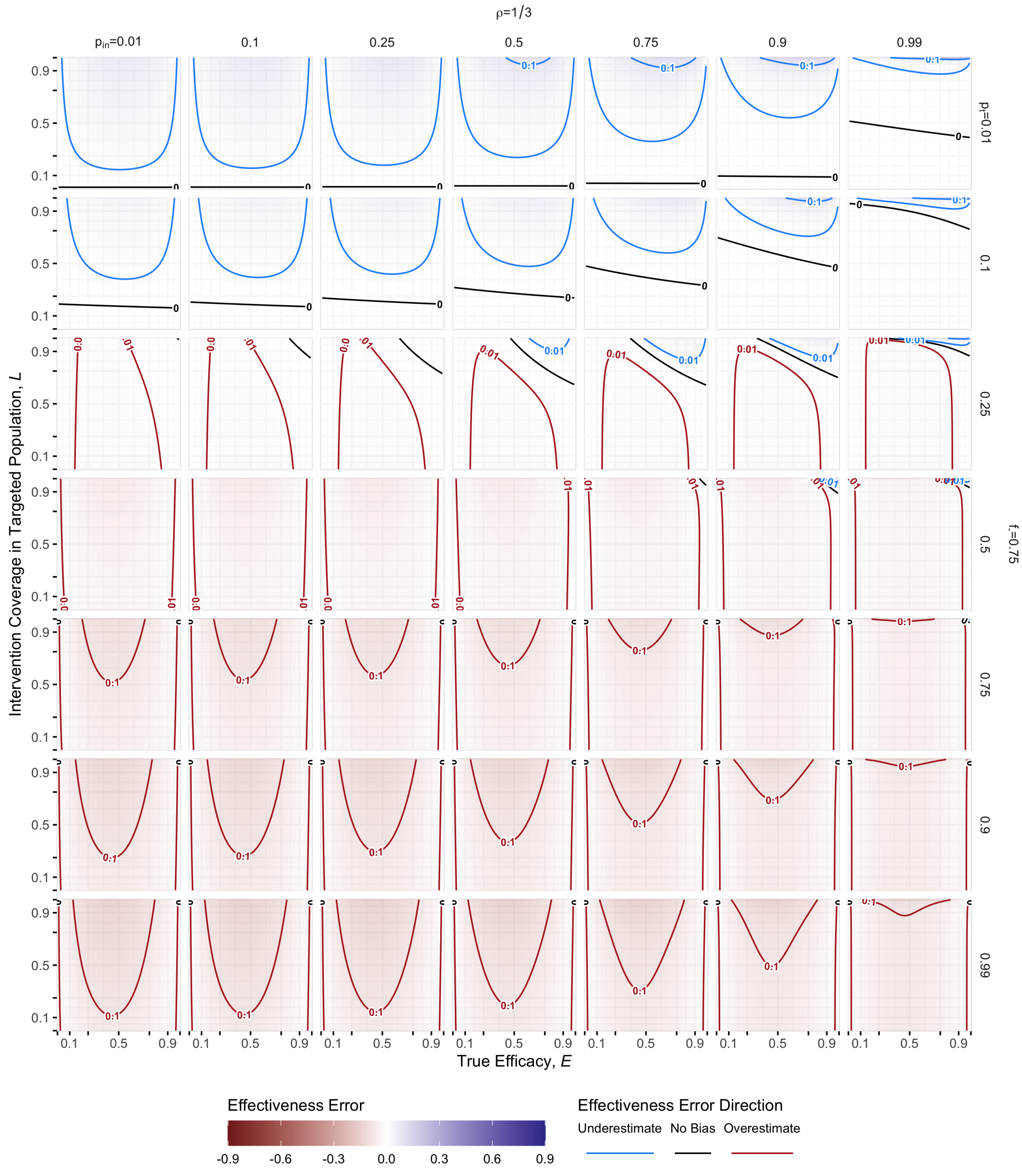

Figure S6: **General Bias Sensitivity. 5 of 9**: Recall, $\rho = \frac{\lambda}{B+1}$ is the expected ratio of secondary to primary recuits; $p_t = R''/\lambda$ is the expected fraction of secondary recruits that test positive when no intervention is present; and $f_- = \frac{B}{B+1}$ is the expect fraction of primary recruits that are test-negative. *In this panel,* $\rho = 1/3$ *and* $f_- = 0.75$.

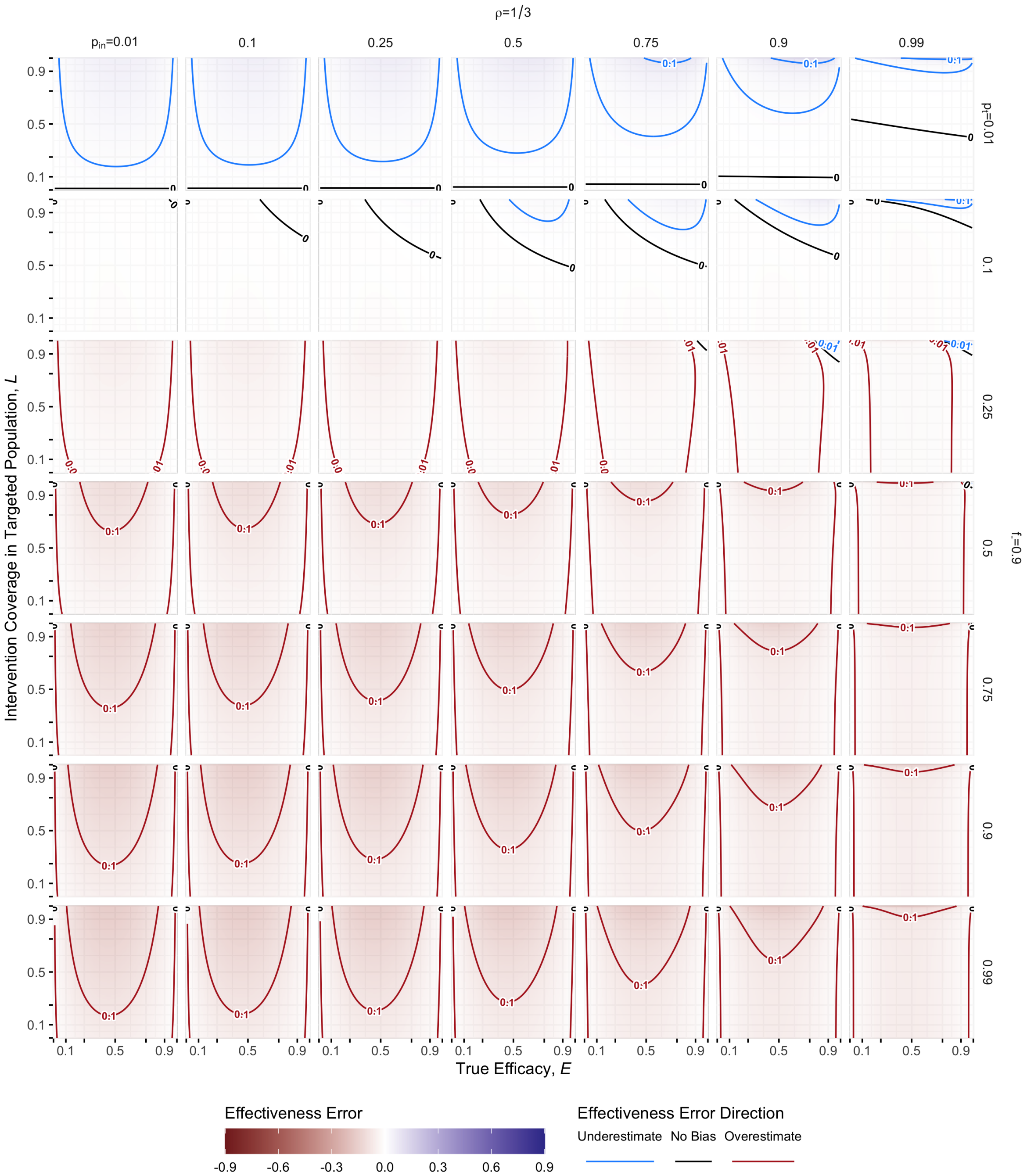

Figure S7: **General Bias Sensitivity. 6 of 9**: Recall, $\rho = \frac{\lambda}{B+1}$ is the expected ratio of secondary to primary recruits; $p_t = R''/\lambda$ is the expected fraction of secondary recruits that test positive when no intervention is present; and $f_- = \frac{B}{B+1}$ is the expect fraction of primary recruits that are test-negative. *In this panel,* $\rho = 1/3$ *and* $f_- = 0.9$.

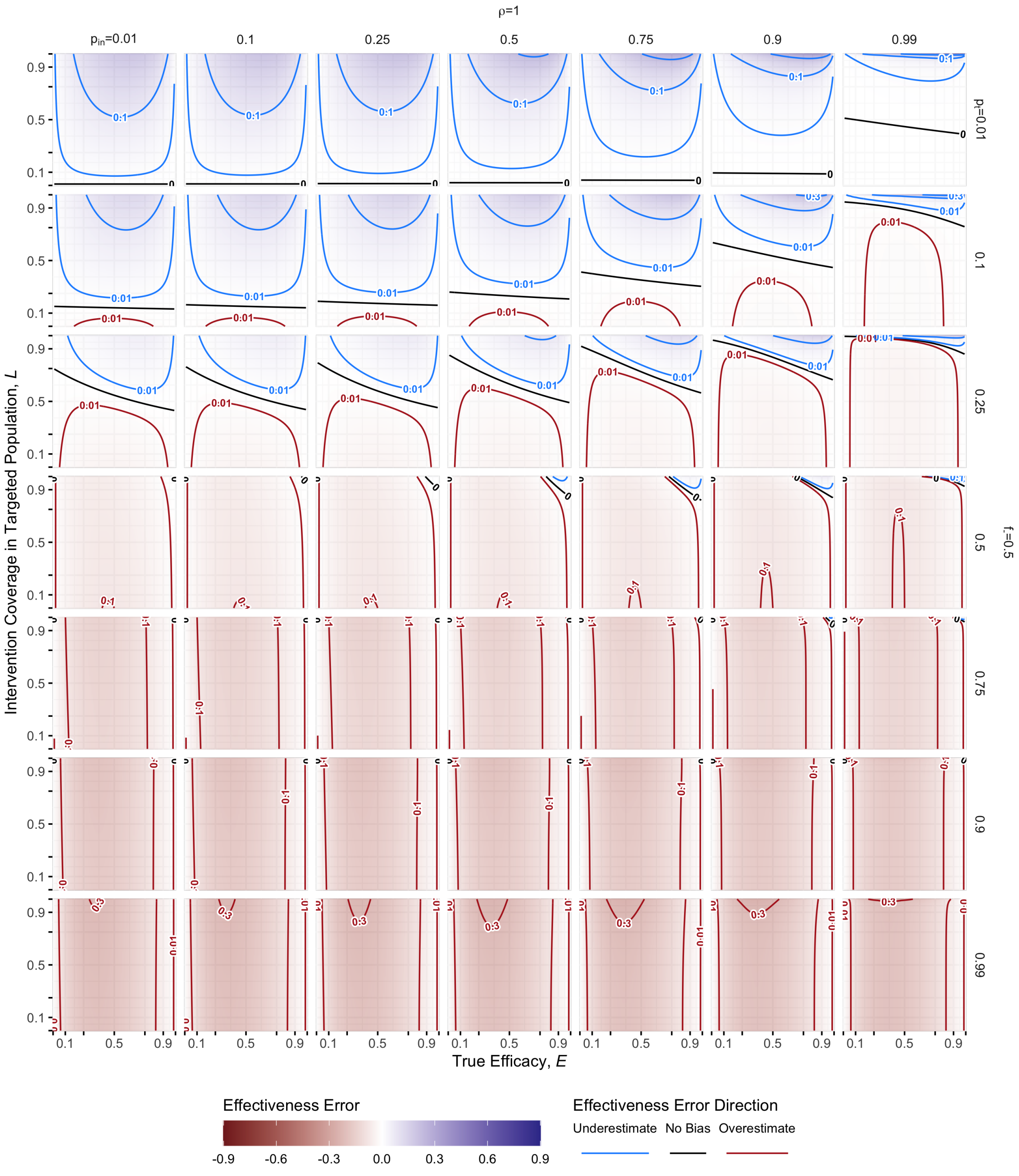

Figure S8: **General Bias Sensitivity. 7 of 9**: Recall, $\rho = \frac{\lambda}{B+1}$ is the expected ratio of secondary to primary recruits; $p_t = R''/\lambda$ is the expected fraction of secondary recruits that test positive when no intervention is present; and $f_- = \frac{B}{B+1}$ is the expect fraction of primary recruits that are test-negative. *In this panel,* $\rho = 1$ *and* $f_- = 0.5$.

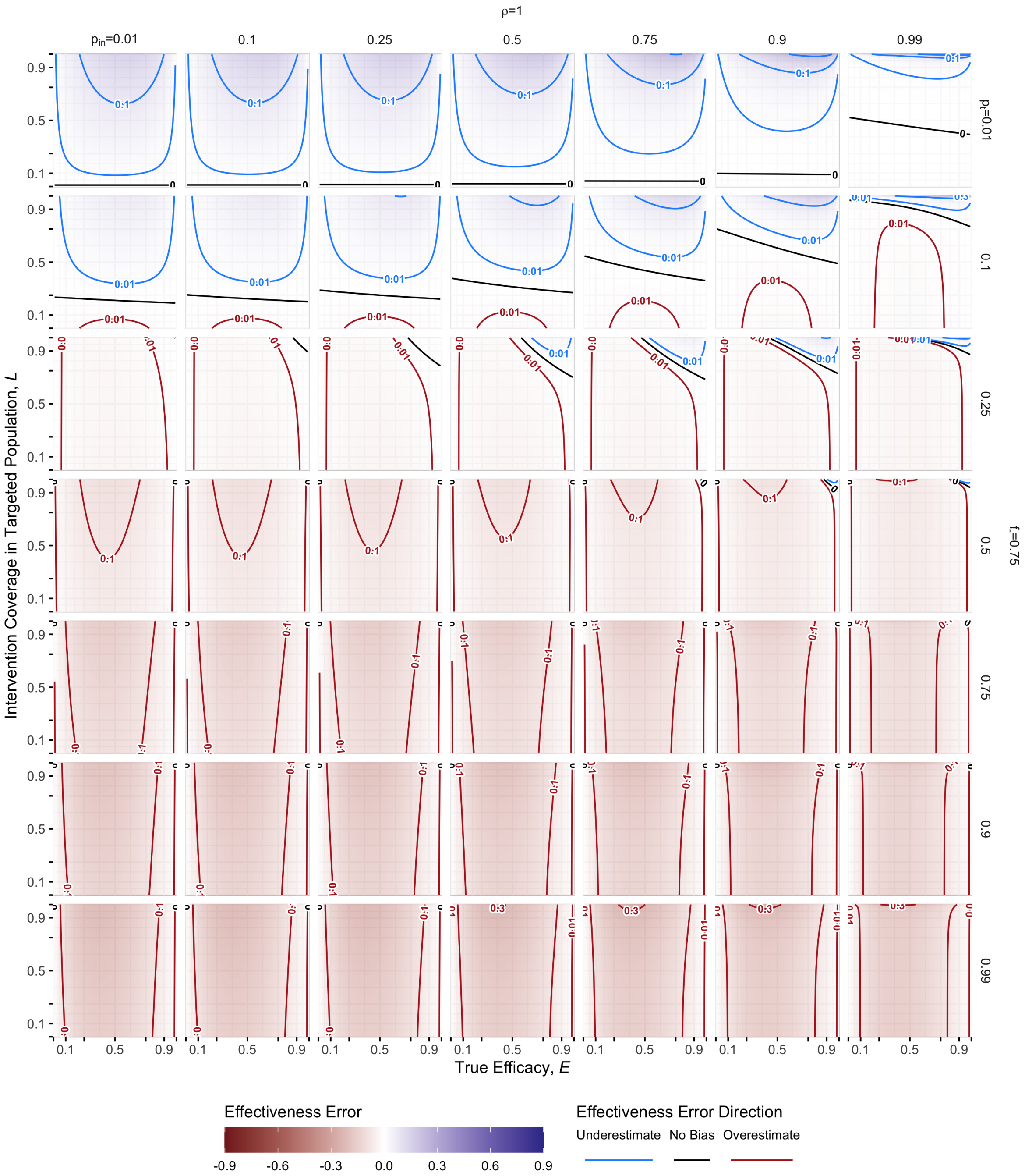

Figure S9: **General Bias Sensitivity. 8 of 9**: Recall, $\rho = \frac{\lambda}{B+1}$ is the expected ratio of secondary to primary recuits; $p_t = R''/\lambda$ is the expected fraction of secondary recruits that test positive when no intervention is present; and $f_- = \frac{B}{B+1}$ is the expect fraction of primary recruits that are test-negative. *In this panel,* $\rho = 1$ *and* $f_- = 0.75$.

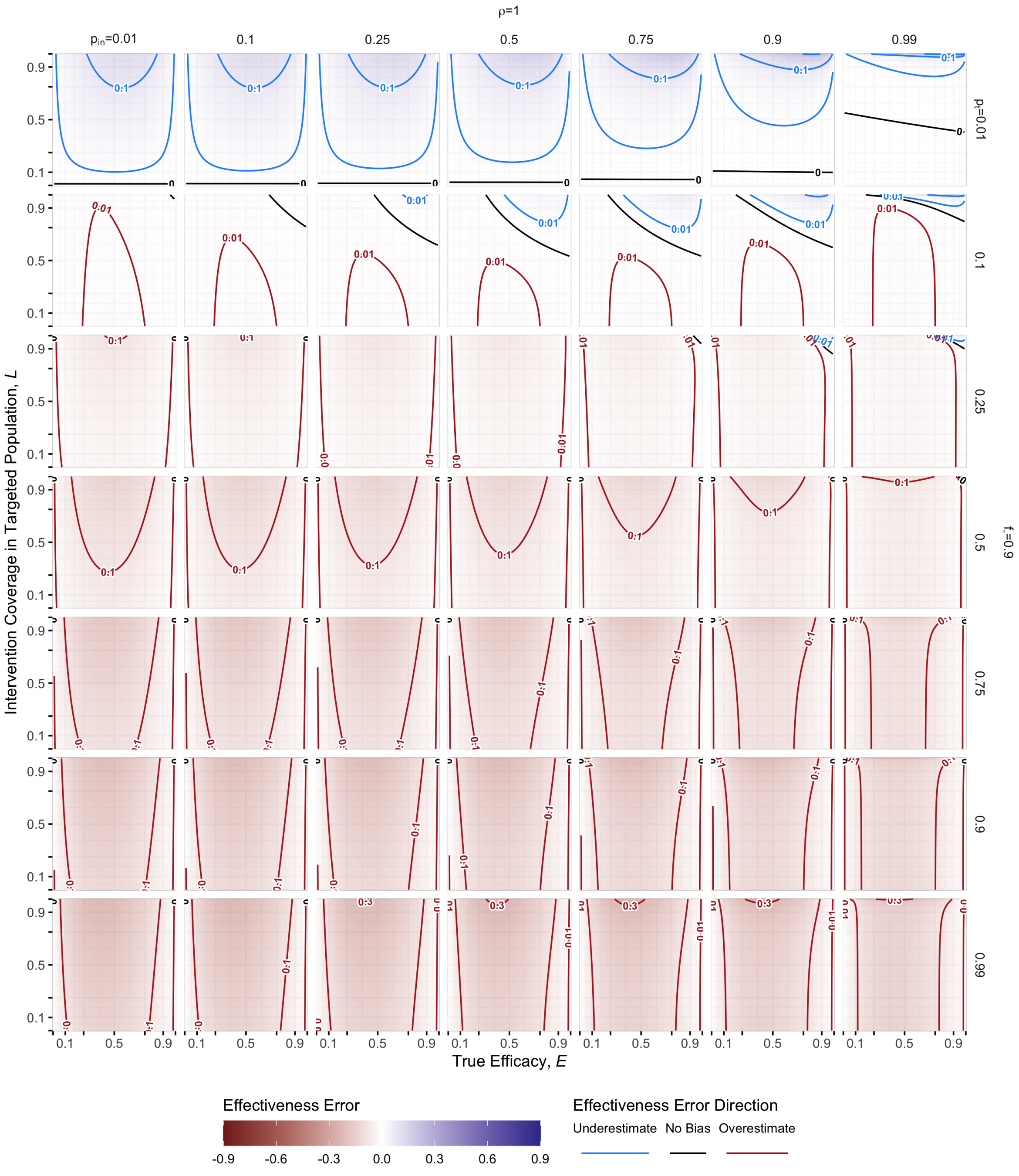

Figure S10: **General Bias Sensitivity.**: Recall, $\rho = \frac{\lambda}{B+1}$ is the expected ratio of secondary to primary recruits; $p_t = R''/\lambda$ is the expected fraction of secondary recruits that test positive when no intervention is present; and $f_- = \frac{B}{B+1}$ is the expect fraction of primary recruits that are test-negative. *In this panel,* $\rho = 1$ *and* $f_- = 0.9$.

#### S6.2 Attempting to Limit to Primary Recruitment Only

An alternative approach to controlling the bias is to restrict to primary recruitment only. If we assume that the study excludes secondary recruitment perfectly for test-positives (*e.g.* because they are extensively monitored) but incompletely excludes secondary recruitment for test-negatives (*e.g.* because data for them is incomplete) then the full estimator equation:

$$\hat{E} = 1 - (1-E)\left[1 + \frac{E\frac{p_t\rho}{1-f_-}}{1+\frac{p_t\rho}{1-f_-}(1-LE)}\frac{L(1-p_{\text{in}})}{1-Lp_{\text{in}}}\right]^{-1}\frac{1+\frac{1-Lp_{\text{in}}-LEp_{\text{in}}+L^2Ep_{\text{in}}}{1-Lp_{\text{in}}-LEp_{\text{in}}+L^2Ep_{\text{in}}^2}\frac{\rho}{f_-}(1-p_t)}{1+\frac{1-LE}{1-LEp_{\text{in}}}\frac{\rho}{f_-}(1-p_t(1-E))}$$

will lose the test-positive bias contribution, because it goes to 1 (note that for any non-zero $p_t$, this term is less than 1):

$$\left[1 + \frac{E\frac{0\rho}{1-f_-}}{1+\frac{0\rho}{1-f_-}(1-LE)}\frac{L(1-p_{\text{in}})}{1-Lp_{\text{in}}}\right]^{-1} = 1 \tag{S33}$$

So the overall bias becomes:

$$\hat{E} = 1 - (1-E)\frac{1+\frac{1-Lp_{\text{in}}-LEp_{\text{in}}+L^2Ep_{\text{in}}}{1-Lp_{\text{in}}-LEp_{\text{in}}+L^2Ep_{\text{in}}^2}\frac{\beta\rho}{f_-}(1-p_t)}{1+\frac{1-LE}{1-LEp_{\text{in}}}\frac{\beta\rho}{f_-}(1-p_t(1-E))} \tag{S34}$$

where $\beta \in (0,1)$ is the exclusion failure probability; $\beta = 0$ is perfect exclusion of secondary test-negatives (in which case the test-negative term also reduces to 1) while $\beta = 1$ implies that no secondary test-negatives are excluded. Note that this factor is simply reducing $\rho$ in the test-negative odds term. Thus, we can drop $\beta$ and instead reduce the range we consider for $\rho$. The error expression for this scenario is:

$$\begin{aligned} E - \hat{E} &= E - 1 + (1-E)\frac{1+\frac{1-Lp_{\text{in}}-LEp_{\text{in}}+L^2Ep_{\text{in}}}{1-Lp_{\text{in}}-LEp_{\text{in}}+L^2Ep_{\text{in}}^2}\frac{\rho}{f_-}(1-p_t)}{1+\frac{1-LE}{1-LEp_{\text{in}}}\frac{\rho}{f_-}(1-p_t(1-E))} \\ &= (1-E)\left[\frac{1+\frac{1-Lp_{\text{in}}-LEp_{\text{in}}+L^2Ep_{\text{in}}}{1-Lp_{\text{in}}-LEp_{\text{in}}+L^2Ep_{\text{in}}^2}\frac{\rho}{f_-}(1-p_t)}{1+\frac{1-LE}{1-LEp_{\text{in}}}\frac{\rho}{f_-}(1-p_t(1-E))} - 1\right] \end{aligned} \tag{S35}$$

For an unbiased estimate, the first term in the square brackets would need to be 1, which would imply:

$$
\begin{aligned}
1 + \frac{1 - Lp_{\text{in}} - LEp_{\text{in}} + L^2Ep_{\text{in}}}{1 - Lp_{\text{in}} - LEp_{\text{in}} + L^2Ep_{\text{in}}^2}\frac{\rho}{f_-}(1-p_t) &= 1 + \frac{1-LE}{1-LEp_{\text{in}}}\frac{\rho}{f_-}(1-p_t(1-E)) \\
\frac{1 - Lp_{\text{in}} - LEp_{\text{in}} + L^2Ep_{\text{in}}}{1 - Lp_{\text{in}} - LEp_{\text{in}} + L^2Ep_{\text{in}}^2}\frac{\rho}{f_-}(1-p_t) &= \frac{1-LE}{1-LEp_{\text{in}}}\frac{\rho}{f_-}(1-p_t(1-E)) \\
\frac{1 - Lp_{\text{in}} - LEp_{\text{in}} + L^2Ep_{\text{in}}}{1 - Lp_{\text{in}}}(1-p_t) &= (1-LE)(1-p_t(1-E)) \\
(1 - Lp_{\text{in}} - LEp_{\text{in}} + L^2Ep_{\text{in}})(1-p_t) &= (1-LE)(1-Lp_{\text{in}})(1-p_t(1-E)) \\
(1 - LEp_{\text{in}} - Lp_{\text{in}}(1-LE))(1-p_t) &= \ldots \\
(1 - LEp_{\text{in}} + LE - LE - Lp_{\text{in}}(1-LE))(1-p_t) &= \ldots \\
((1-Lp_{\text{in}})(1-LE) + LE(1-p_{\text{in}}))(1-p_t) &= \ldots \\
(1-Lp_{\text{in}})(1-LE)(1-p_t) + LE(1-p_{\text{in}})(1-p_t) &= (1-LE)(1-Lp_{\text{in}})(1-p_t) + (1-LE)(1-Lp_{\text{in}})Ep_t \\
LE(1-p_{\text{in}})(1-p_t) &= (1-LE)(1-Lp_{\text{in}})Ep_t \\
L(1-p_{\text{in}})(1-p_t) &= (1-LE)(1-Lp_{\text{in}})p_t
\end{aligned}
\tag{S36}
$$

There is no further reduction to the final line of Eq. S36, thus this term is only equal to 1 for specific combinations of $\{L, E, p_{\text{in}}, p_t\}$. Nor is there a strict direction of inequality. For example, at $L \approx 0$, the left hand side is less than or equal to the right, while at $L \approx 1$ the inequality can be either direction depending on the value of $p_t$. The directions of this inequality determine whether the residual term in Eq. S36 is greater than 1 (*i.e.*, the left hand side is greater than the right) or less than 1 (*vice versa*).

Defining this residual term as $Y$ for the moment, and term corresponding to the test-positives as $X$ (which recall is $X \leq 1$), we can consider the magnitude of the error excluding only the secondary test-positives versus keeping all the secondary recruiting by:

$$
\frac{|E - \hat{E}|}{|E - \hat{E}^*|} = \frac{(1-E)|YX - 1|}{(1-E)|Y-1|} = \frac{|YX-1|}{|Y-1|} \tag{S37}
$$

For $Y \leq 1$, we know $XY \leq Y \leq 1$ and thus $XY - 1 \leq Y - 1 \leq 0$. This implies reduced (or at least the same) error magnitude whenever $Y \leq 1$ holds, *i.e.* the left hand side of Eq. S36 is less than or equal to the right. Decreasing $E$ always increases the right hand side without effecting the left, making the constraint harder to satisfy and the least true efficacy we considered was $E = 0$. Increasing $p_t$ always decreases the left hand side while increasing the right, so the smallest $p_t$ is also the most restrictive condition to meet this criterion. In the main text, the smallest value we considered was $p_t \approx 0.07$. Under these circumstances:

$$
\frac{L(1-p_{\text{in}})}{1-Lp_{\text{in}}} \overset{?}{\leq} \frac{p_t}{(1-p_t)} \approx 0.07 \tag{S38}
$$

This only true for low $L$ and high $p_{\text{in}}$ combinations, outside what we considered in the main text.

For $Y > 1$, some reduction in $XY$ due the $X \leq 1$ term reduces bias magnitude, namely if

$$
XY \geq 1 - (Y-1) \implies X \geq \frac{2-Y}{Y} \tag{S39}
$$

but too much will overshoot and lead to increased bias magnitude (though in the other direction). Of course, $X$ and $Y$ share terms, so

$$X \geq \frac{2-Y}{Y} \implies$$

$$1 + \frac{E\frac{p_t\rho}{1-f_-}}{1+\frac{p_t\rho}{1-f_-}(1-LE)} \frac{L(1-p_{\text{in}})}{1-Lp_{\text{in}}} \leq \frac{\frac{1+\frac{1-Lp_{\text{in}}-LEp_{\text{in}}+L^2Ep_{\text{in}}}{1-Lp_{\text{in}}-LEp_{\text{in}}+L^2Ep_{\text{in}}^2}\frac{\rho}{f_-}(1-p_t)}{1+\frac{1-LE}{1-LEp_{\text{in}}}\frac{\rho}{f_-}(1-p_t(1-E))}}{2-\frac{1+\frac{1-Lp_{\text{in}}-LEp_{\text{in}}+L^2Ep_{\text{in}}}{1-Lp_{\text{in}}-LEp_{\text{in}}+L^2Ep_{\text{in}}^2}\frac{\rho}{f_-}(1-p_t)}{1+\frac{1-LE}{1-LEp_{\text{in}}}\frac{\rho}{f_-}(1-p_t(1-E))}} \tag{S40}$$

$$\begin{aligned}
E-\hat{E} &= E-1+(1-E)\frac{1+\frac{1-Lp_{\text{in}}-LEp_{\text{in}}+L^2Ep_{\text{in}}}{1-Lp_{\text{in}}-LEp_{\text{in}}+L^2Ep_{\text{in}}^2}\frac{\rho}{f_-}(1-p_t)}{1+\frac{1-LE}{1-LEp_{\text{in}}}\frac{\rho}{f_-}(1-p_t(1-E))} \\
&= -(1-E)\left[1-\frac{1+\frac{1-Lp_{\text{in}}-LEp_{\text{in}}+L^2Ep_{\text{in}}}{1-Lp_{\text{in}}-LEp_{\text{in}}+L^2Ep_{\text{in}}^2}\frac{\rho}{f_-}(1-p_t)}{1+\frac{1-LE}{1-LEp_{\text{in}}}\frac{\rho}{f_-}(1-p_t(1-E))}\right] \\
&= -(1-E)\left[\frac{\frac{1-LE}{1-LEp_{\text{in}}}\frac{\rho}{f_-}(1-p_t(1-E))-\frac{1-Lp_{\text{in}}-LEp_{\text{in}}+L^2Ep_{\text{in}}}{1-Lp_{\text{in}}-LEp_{\text{in}}+L^2Ep_{\text{in}}^2}\frac{\rho}{f_-}(1-p_t)}{1+\frac{1-LE}{1-LEp_{\text{in}}}\frac{\rho}{f_-}(1-p_t(1-E))}\right] \\
&= \frac{-(1-E)\frac{\rho}{f_-}}{(1-Lp_{\text{in}})}\frac{(1-Lp_{\text{in}})(1-LE)(1-p_t(1-E))-\left(1-Lp_{\text{in}}-LEp_{\text{in}}+L^2Ep_{\text{in}}\right)(1-p_t)}{(1-LEp_{\text{in}})+(1-LE)\frac{\rho}{f_-}(1-p_t(1-E))} \\
&= \frac{-(1-E)\frac{\rho}{f_-}}{(1-Lp_{\text{in}})}\frac{(1-Lp_{\text{in}}-LE+L^2Ep_{\text{in}})(1-p_t(1-E))-\left(1-Lp_{\text{in}}-LEp_{\text{in}}+L^2Ep_{\text{in}}\right)(1-p_t)}{(1-LEp_{\text{in}})+(1-LE)\frac{\rho}{f_-}(1-p_t(1-E))} \\
&= \frac{-(1-E)\frac{\rho}{f_-}}{(1-Lp_{\text{in}})}\frac{(1-Lp_{\text{in}}-LE+L^2Ep_{\text{in}})Ep_t+-LE(1-p_t)+LEp_{\text{in}}(1-p_t)}{(1-LEp_{\text{in}})+(1-LE)\frac{\rho}{f_-}(1-p_t(1-E))} \\
&= \frac{-(1-E)\frac{\rho}{f_-}}{(1-Lp_{\text{in}})}\frac{(1-Lp_{\text{in}}-LE+L^2Ep_{\text{in}})Ep_t-LE(1-p_t)(1-p_{\text{in}})}{(1-LEp_{\text{in}})+(1-LE)\frac{\rho}{f_-}(1-p_t(1-E))} \\
&= \frac{-(1-E)E\frac{\rho}{f_-}}{(1-Lp_{\text{in}})}\frac{(1-Lp_{\text{in}})(1-LE)p_t-L(1-p_t)(1-p_{\text{in}})}{(1-LEp_{\text{in}})+(1-LE)\frac{\rho}{f_-}(1-p_t(1-E))}
\end{aligned} \tag{S41}$$

Because $p_{\text{in}}$ remains in the equation, in multiple places, the targeted fraction plays an important role in bias. If targeted fraction is high, and for the case of $p_{\text{in}} \to 1$, then we can then recover:

$$\begin{aligned}
\lim_{p_{\text{in}}\to 1} E-\hat{E} &= \frac{-(1-E)E\frac{\rho}{f_-}}{(1-L)}\frac{(1-L)(1-LE)p_t-L(1-p_t)(1-1)}{(1-LE)+(1-LE)\frac{\rho}{f_-}(1-p_t(1-E))} \\
&= -(1-E)E\frac{\rho}{f_-}\frac{p_t}{1+\frac{\rho}{f_-}(1-p_t(1-E))} \\
&= -(1-E)E\frac{p_t}{1+\frac{f_-}{\rho}-p_t(1-E))}
\end{aligned} \tag{S42}$$

which is the same error as if we had just been able to control $p_{\text{in}} \to 1$. This suggests that if the study could mostly manage heterogeneity in the recruited population, but likely could not exclude secondary test-negatives, then including all secondary cases does not affect the magnitude of bias.

Figures S11-S19 show the response of the estimator to varying factors when secondary test-positives are excluded. Each plot shows $p_{\text{in}} \in \{0.01, 0.1, 0.25, 0.5, 0.75, 0.9, 1\}$ (columns) and $p_t \in$

$\{0.01, 0.1, 0.25, 0.5, 0.75, 0.9, 1\}$ (rows). Each plot shows one of the combinations of $\rho \in \{1/9, 1/3, 1\}$ and $f_- \in \{0.5, 0.75, 0.9\}$; $\rho$ is indicated at the top of each plot, $f_-$ on the right side.

In general, these plots show the same trends as Figures S2-S10, with lower bias magnitude and tendency to shift towards underestimation.

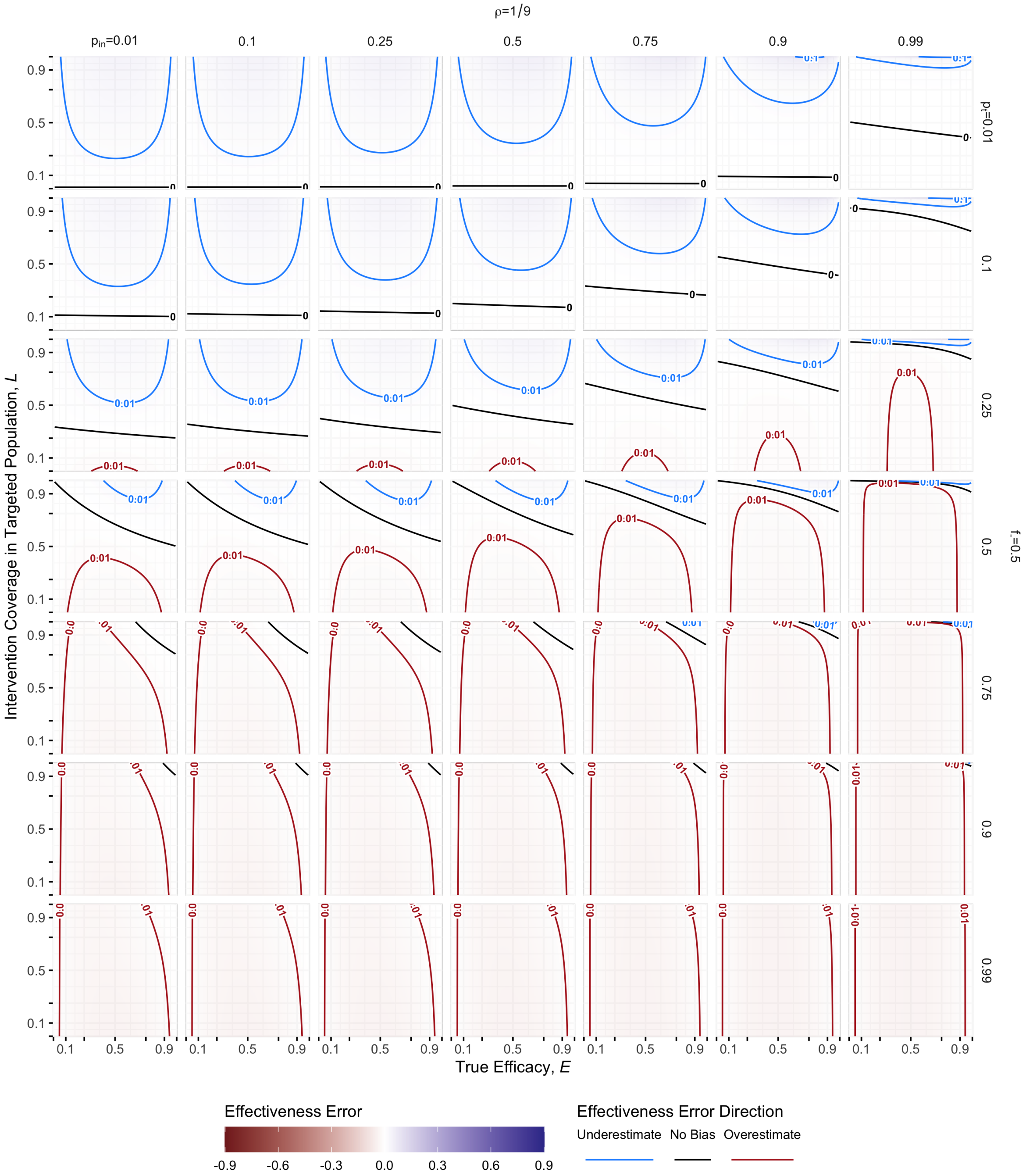

Figure S11: **Bias Sensitivity Without Secondary Test-Positives, 1 of 9**: These series of plots show sensitivity of the TND estimator which excludes secondary test-positives to all of the model parameters. For each plot, the rightmost column corresponds to very high (99%) targeted fraction, which indicates the minimal bias surface when the study manages to maximize targeted fraction. Recall, $\rho = \frac{\lambda}{B+1}$ is the expected ratio of secondary to primary recuits; $p_t = R''/\lambda$ is the expected fraction of secondary recruits that test positive when no intervention is present; and $f_- = \frac{B}{B+1}$ is the expect fraction of primary recruits that are test-negative. *In this panel,* $\rho = 1/9$ *and* $f_- = 0.5$.

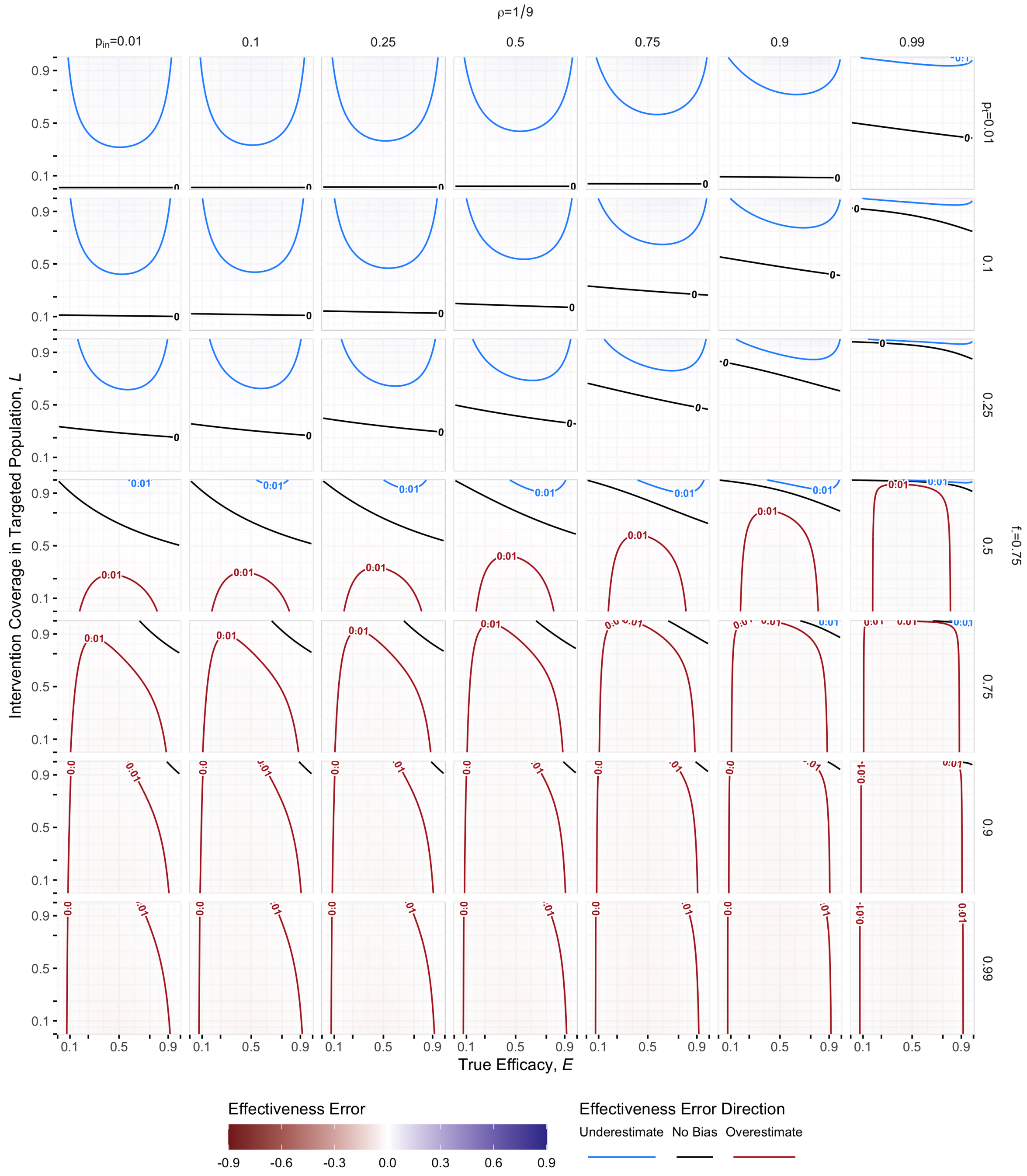

Figure S12: **Bias Sensitivity Without Secondary Test-Positives. 2 of 9**: Recall, $\rho = \frac{\lambda}{B+1}$ is the expected ratio of secondary to primary recuits; $p_t = R''/\lambda$ is the expected fraction of secondary recruits that test positive when no intervention is present; and $f_{-} = \frac{B}{B+1}$ is the expect fraction of primary recruits that are test-negative. *In this panel,* $\rho = 1/9$ *and* $f_{-} = 0.75$.

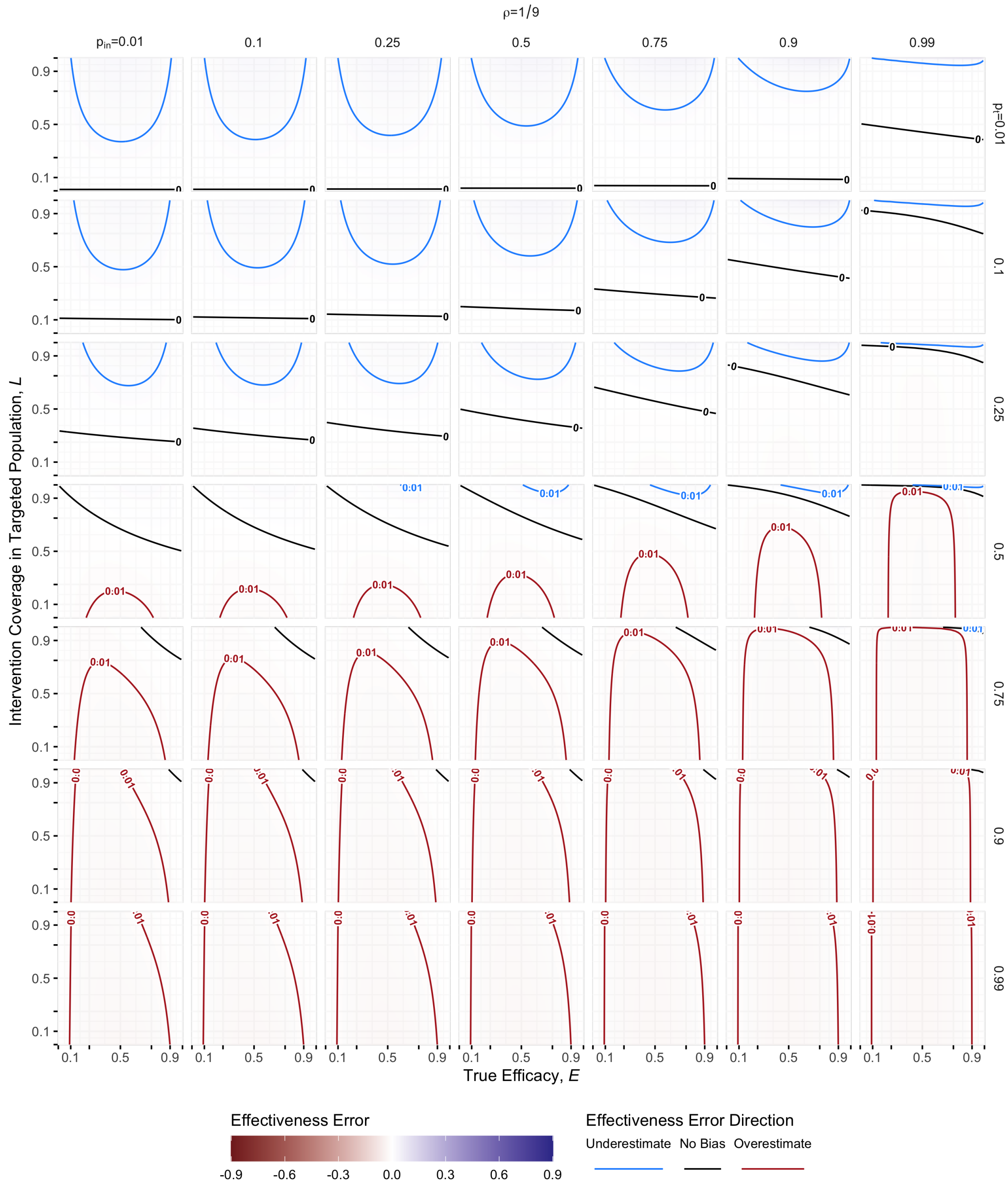

Figure S13: **Bias Sensitivity Without Secondary Test-Positives. 3 of 9**: Recall, $\rho = \frac{\lambda}{B+1}$ is the expected ratio of secondary to primary recuits; $p_t = R''/\lambda$ is the expected fraction of secondary recruits that test positive when no intervention is present; and $f_- = \frac{B}{B+1}$ is the expect fraction of primary recruits that are test-negative. *In this panel,* $\rho = 1/9$ *and* $f_- = 0.9$*.*

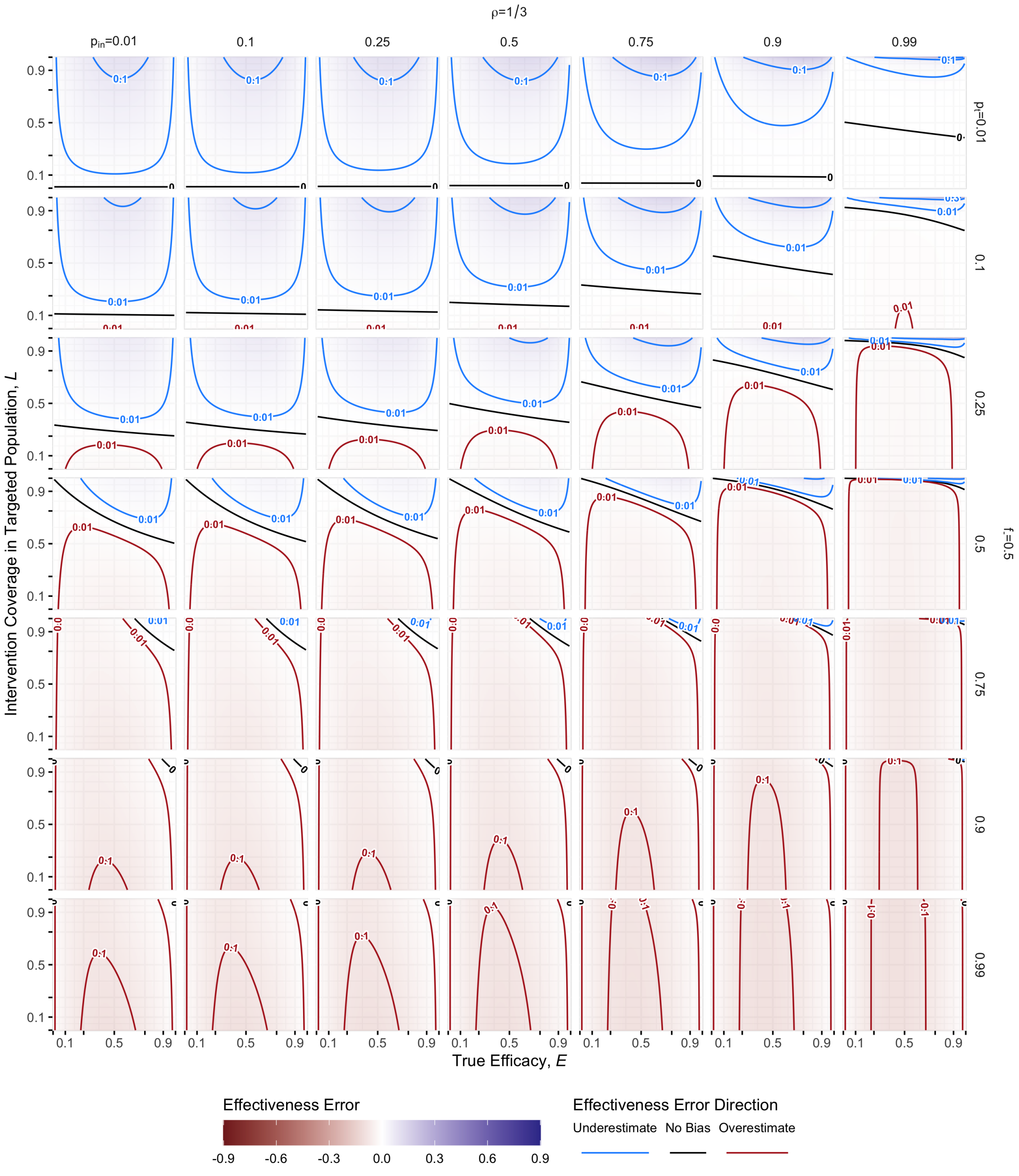

Figure S14: **Bias Sensitivity Without Secondary Test-Positives. 4 of 9**: Recall, $\rho = \frac{\lambda}{B+1}$ is the expected ratio of secondary to primary recuits; $p_t = R''/\lambda$ is the expected fraction of secondary recruits that test positive when no intervention is present; and $f_- = \frac{B}{B+1}$ is the expect fraction of primary recruits that are test-negative. *In this panel,* $\rho = 1/3$ *and* $f_- = 0.5$.

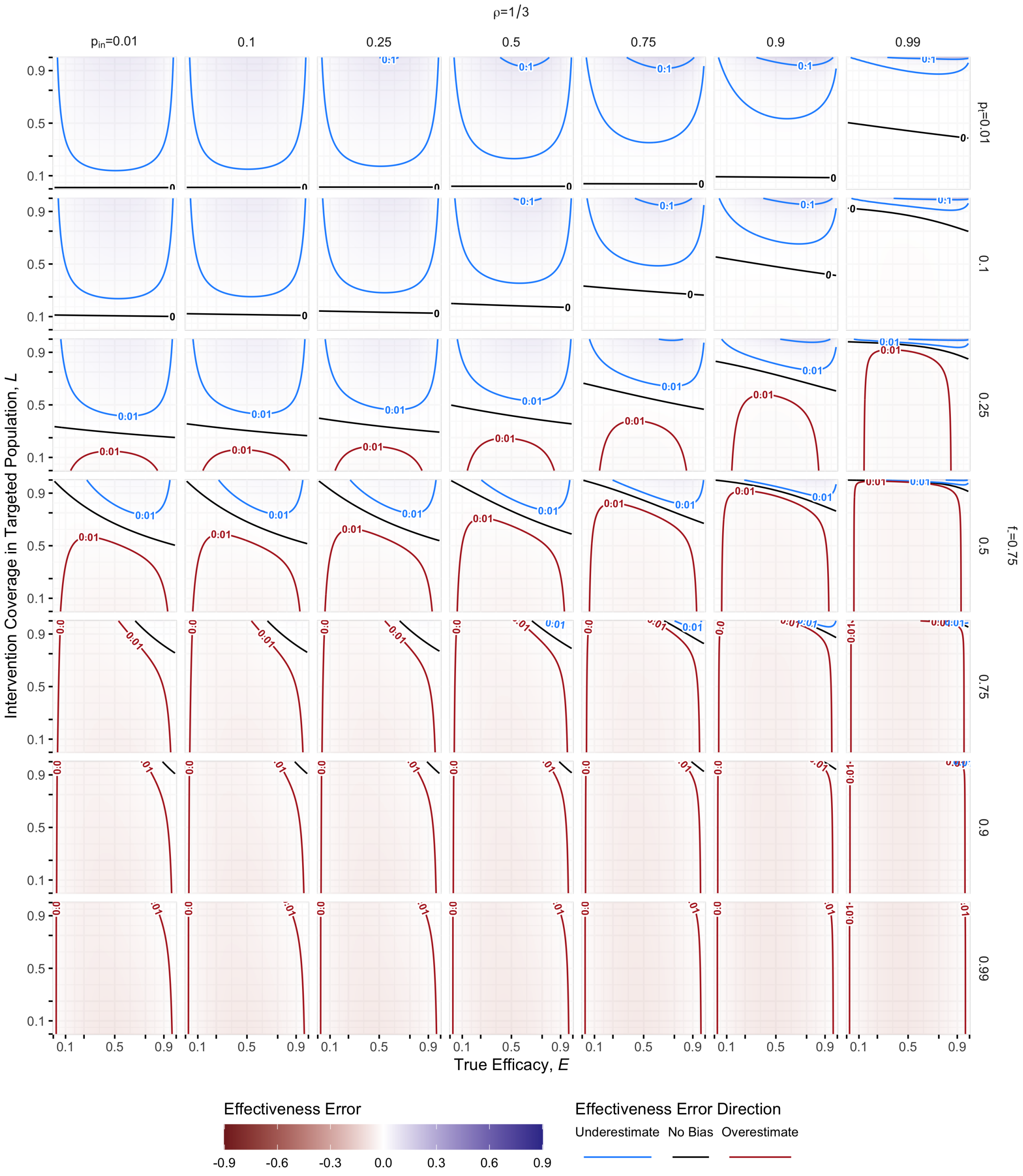

Figure S15: **Bias Sensitivity Without Secondary Test-Positives. 5 of 9**: Recall, $\rho = \frac{\lambda}{B+1}$ is the expected ratio of secondary to primary recuits; $p_t = R''/\lambda$ is the expected fraction of secondary recruits that test positive when no intervention is present; and $f_- = \frac{B}{B+1}$ is the expect fraction of primary recruits that are test-negative. *In this panel,* $\rho = 1/3$ *and* $f_- = 0.75$.

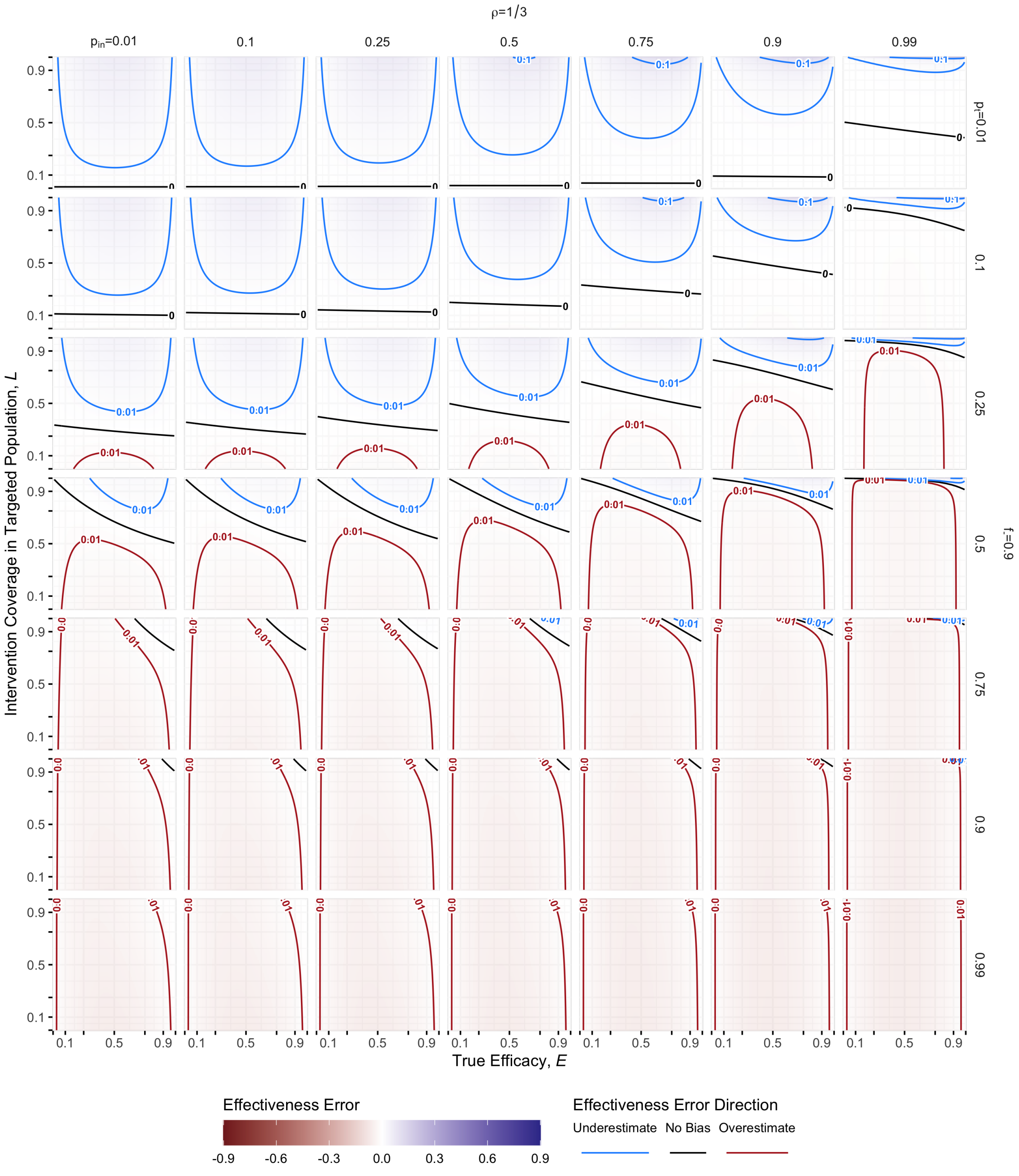

Figure S16: **Bias Sensitivity Without Secondary Test-Positives. 6 of 9**: Recall, $\rho = \frac{\lambda}{B+1}$ is the expected ratio of secondary to primary recuits; $p_t = R''/\lambda$ is the expected fraction of secondary recruits that test positive when no intervention is present; and $f_- = \frac{B}{B+1}$ is the expect fraction of primary recruits that are test-negative. *In this panel,* $\rho = 1/3$ *and* $f_- = 0.9$.

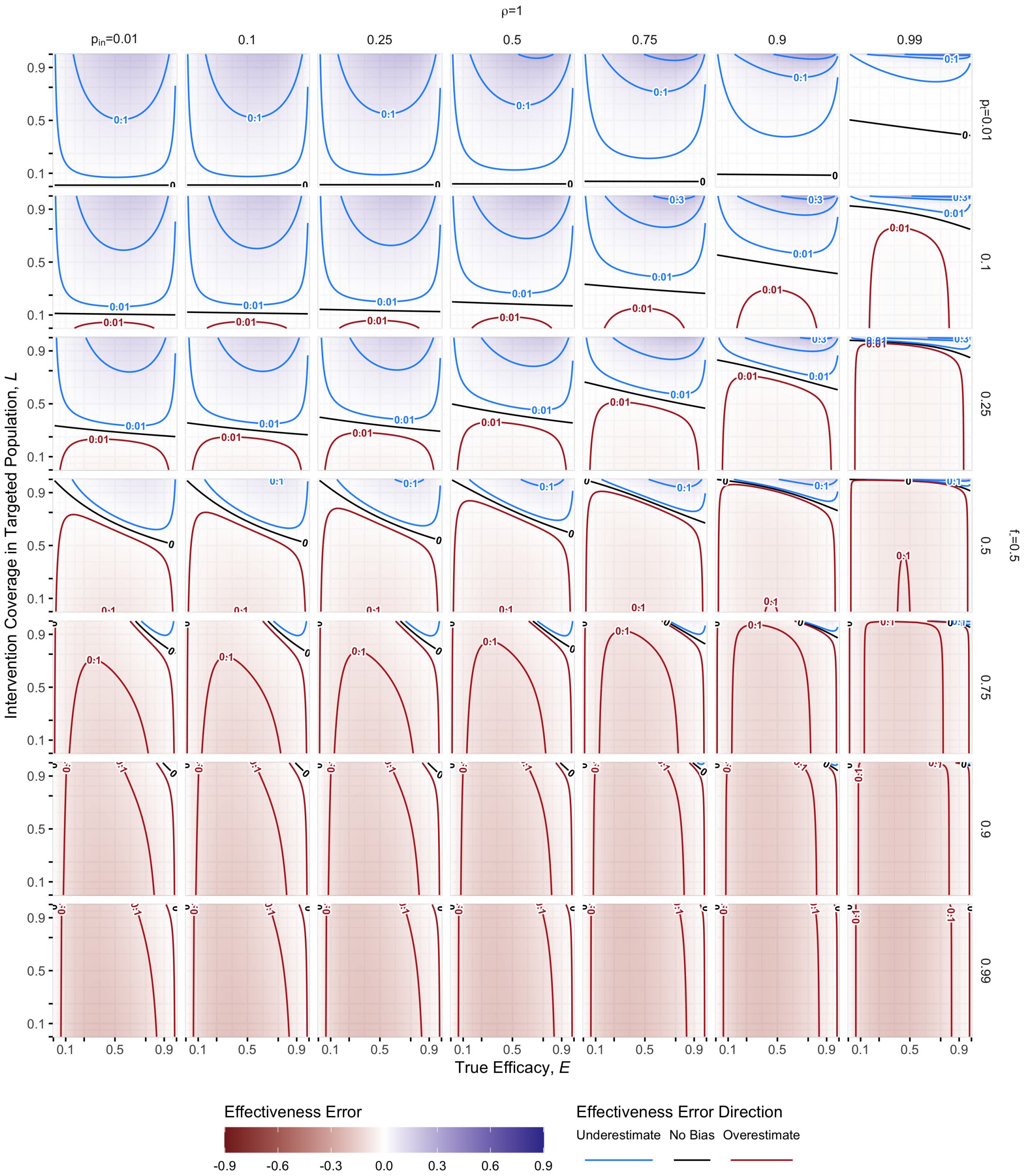

Figure S17: **Bias Sensitivity Without Secondary Test-Positives. 7 of 9**: Recall, $\rho = \frac{\lambda}{B+1}$ is the expected ratio of secondary to primary recuits; $p_t = R''/\lambda$ is the expected fraction of secondary recruits that test positive when no intervention is present; and $f_- = \frac{B}{B+1}$ is the expect fraction of primary recruits that are test-negative. *In this panel,* $\rho = 1$ *and* $f_- = 0.5$.

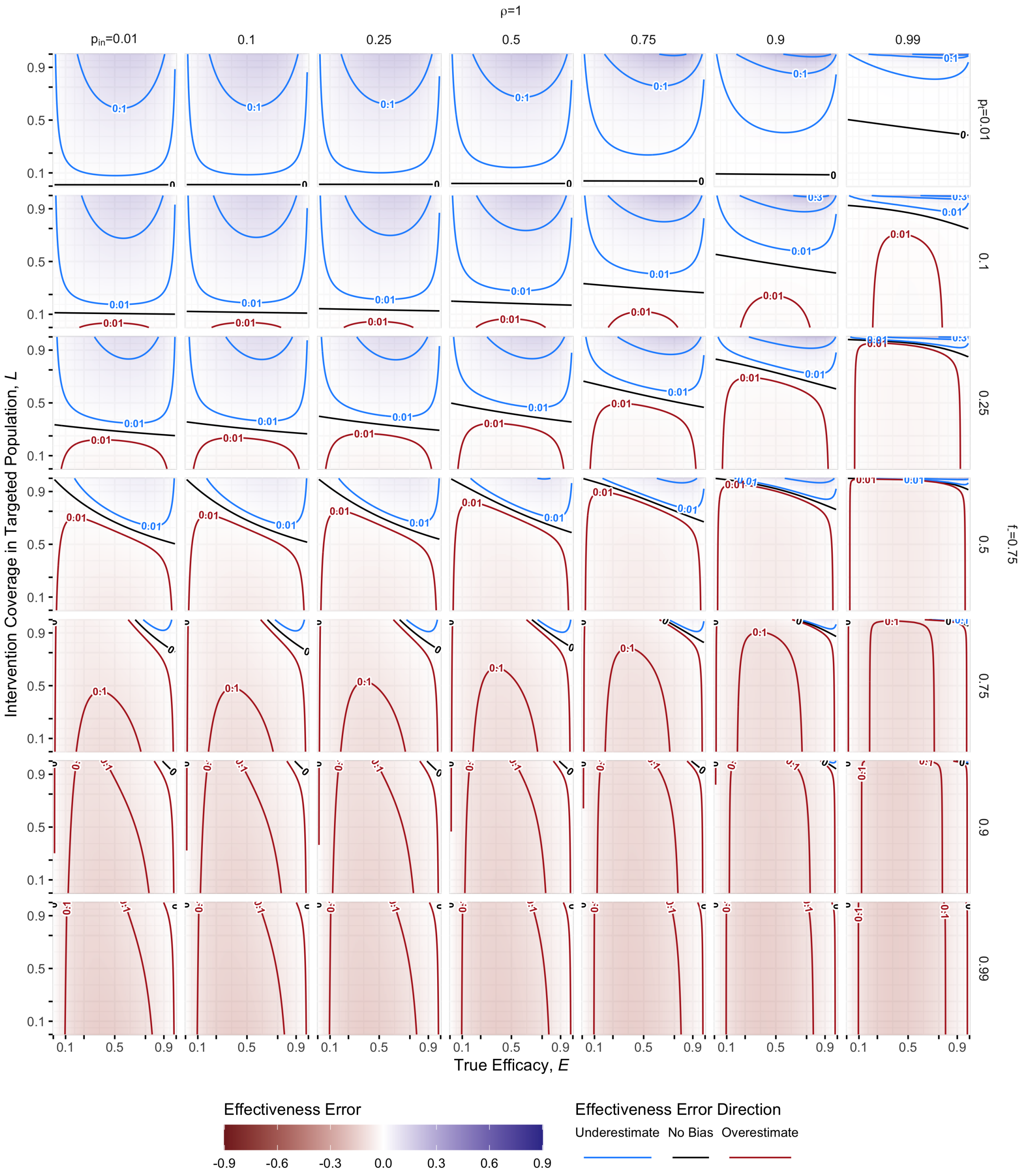

Figure S18: **Bias Sensitivity Without Secondary Test-Positives. 8 of 9**: Recall, $\rho = \frac{\lambda}{B+1}$ is the expected ratio of secondary to primary recuits; $p_t = R''/\lambda$ is the expected fraction of secondary recruits that test positive when no intervention is present; and $f_- = \frac{B}{B+1}$ is the expect fraction of primary recruits that are test-negative. *In this panel,* $\rho = 1$ *and* $f_- = 0.75$.

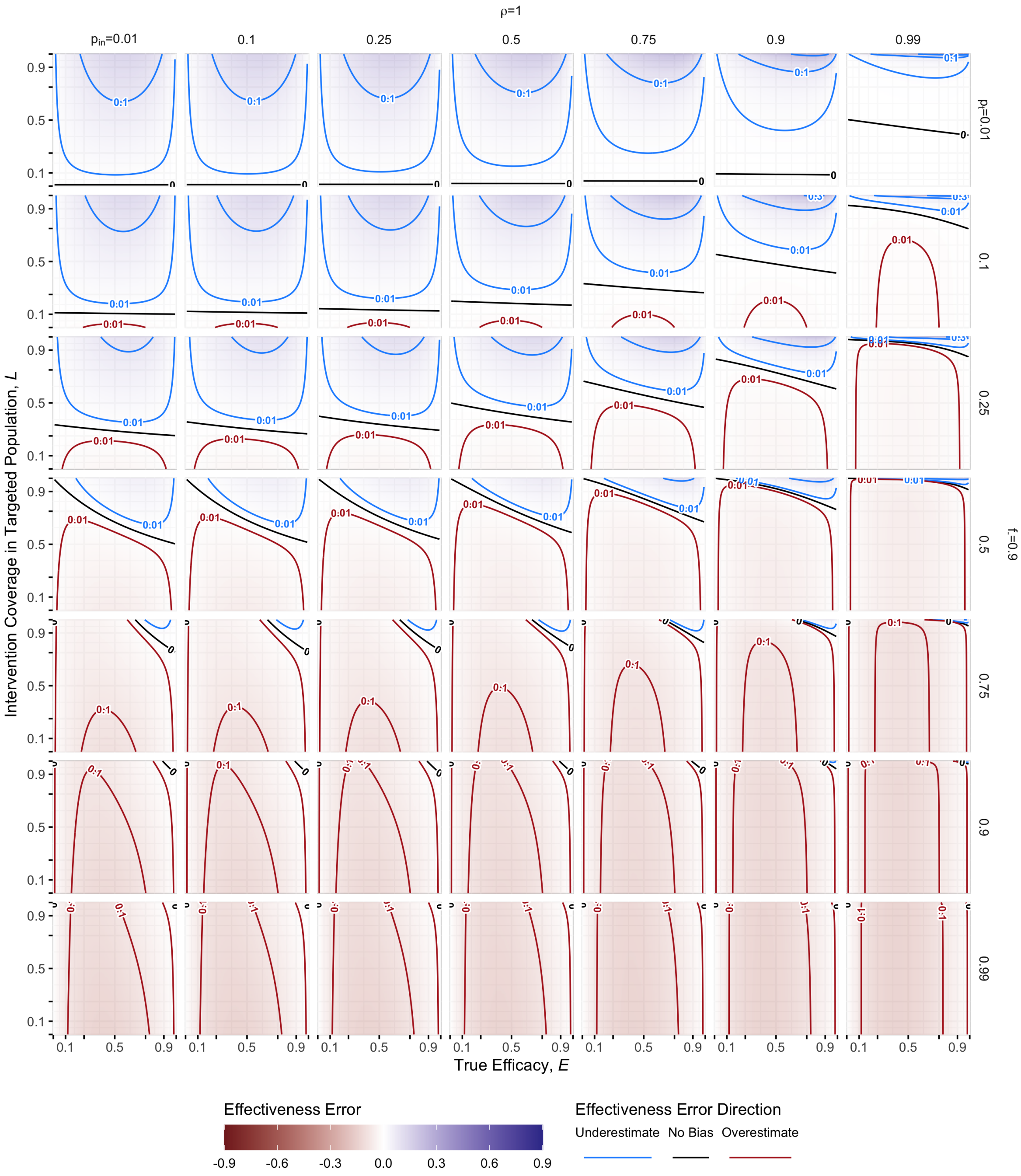

Figure S19: **Bias Sensitivity Without Secondary Test-Positives.**: Recall, $\rho = \frac{\lambda}{B+1}$ is the expected ratio of secondary to primary recuits; $p_t = R''/\lambda$ is the expected fraction of secondary recruits that test positive when no intervention is present; and $f_- = \frac{B}{B+1}$ is the expect fraction of primary recruits that are test-negative. *In this panel,* $\rho = 1$ *and* $f_- = 0.9$.

### S7 Calculation of Coverage, $L$, and Targeted Fraction, $p_{\text{in}}$

In addition to parameters associated with epidemiology and response activities ($B$, $\lambda$, and $R''$), the values of $L$ and $p_{\text{in}}$ are required to determine what level of bias may be present. Our model proposes a population where there are three groups, non-targeted, and targeted individuals that receive the intervention or not. If we have an estimate for the size of the total recruitable population, we can use measures taken during the intervention distribution to estimate these values. Alternatively, additional data could be collected when testing recruits to estimate these values. The estimates proposed hereafter are not definitive; in addition to assuming our model of heterogeneity is a sufficiently useful approximation, they make further strong assumptions about behaviour around intervention uptake. However, as estimates they are potentially informative about the limits of $p_{\text{in}}$ and $L$ within the model framework we have proposed.

#### S7.1 Measures During Intervention Distribution

During distribution of the intervention, one could count the number of people receiving the intervention ($V$) and the number ineligible ($U*$). From those values we can compute the crude minimum and maximum values of $L$ and $p_{\text{in}}$ (where $\max(L)$ corresponds to $\min(p_{\text{in}})$, and *vice versa*).

$$
\begin{aligned}
L &\in \left(\frac{V}{T}, \frac{V}{V+U*}\right) \\
p_{\text{in}} &\in \left(0, 1 - \frac{V+U*}{T}\right)
\end{aligned}
\tag{S43}
$$

In the model, we make no assumptions of how non-intervention occurs in the targeted population, just that it occurs randomly within that group. Thus the ineligible count represents the minimum non-intervention amongst the targeted population (the upper limit of $L$); there may be other sources (*e.g.*, targeted individuals are unavailable on the day offered). Potentially, when distributing the intervention, targeted individuals could be asked about members of their household, neighbors, *etc.* that wanted to get the intervention, but were unable to do so, but this number would also have many uncertainties (*e.g.* duplicate reporting, reporting individuals that do receive the intervention at a different time or place).

If the study intervention has multiple steps (*e.g.* a two-dose vaccine, repeat application of vector-control insecticides), then the decrease in coverage between steps could be informative about the targeted fraction, $p_{\text{in}}$. If we assume not receiving the intervention is due to a mix of short-term (*e.g.* ill that day) and long-term (*e.g.* too young to be eligible) effects, then we can potentially further constrain $L$ and $p_{\text{in}}$. In the following we assume that: i) long-term ineligibles only present themselves at the first step (though they may also not), ii) short-term ineligibles present at the same rate in the subsequent steps, and iii) individuals that did not present at earlier steps will also not present later. If we apply these assumptions to a two-step intervention, and we call unobserved long-term ineligibles $I_0$, the long-term ineligibles that appear initially are $I_1$, and the intervention recipients ($V$) and short-term ineligibles that present ($U^*$) or not ($U$) at each stage ($V_1, V_2, U*_1, U*_2, U_1, U_2$), then the following relations hold.

We have six observed pieces of information: the total population ($T$), the number given the intervention versus short term ineligible at both steps ($V_1, V_2, U*_1, U*_2$), and the number of long-term ineligibles at the first step ($I_1$).

We know that at the second intervention step, we have only people that got vaccinated in the previous step, no new long term ineligibles, and the breakdown of short term ineligibles versus those that receive the second step. So:

$$V_1 = U_2 + U_2^* + V_2 \implies U_2 = V_1 - V_2 - U_2^* \tag{S44}$$

If we assume that the division between presenting versus not presenting for short-term ineligibles is the same for both steps, then:

$$\begin{aligned}
\frac{U_2^*}{U_2 + U_2^*} = \frac{U_1^*}{U_1 + U_1{*}} &\implies \frac{U_2^*}{U_2 + U_2^*}\left(U_1 + U_1{*}\right) = U_1^* \\
&\implies \frac{U_2^*}{U_2 + U_2^*} U_1 = U_1^*\left(1 - \frac{U_2^*}{U_2 + U_2^*}\right) \\
&\implies U_1^* \frac{V_1 - V_2 - U_2^*}{U_2^*} = U_1
\end{aligned} \tag{S45}$$

For the first distribution of the intervention, we assume that the ratio of short- and long-term ineligibles presenting is the same as those not presenting:

$$\begin{aligned}
\frac{U_1^*}{U_1^* + I_1} = \frac{U_1}{U_1 + I_0} &\implies \frac{U_1^*}{U_1^* + I_1}\left(U_1 + I_0\right) = U_1 \\
&\implies \frac{U_1^*}{U_1^* + I_1} I_0 = U_1\left(1 - \frac{U_1^*}{U_1^* + I_1}\right) \\
&\implies I_0 = U_1 \frac{I_1}{U_1^*} = I_1 \frac{V_1 - V_2 - U_2^*}{U_2^*}
\end{aligned} \tag{S46}$$

Finally, the pieces must add up to the total population, and therefore:

$$\begin{aligned}
C &= V_1 + U_1^* + I_1 + U_1 + I_0 \\
&= V_1 + U_1^*\left(1 + \frac{V_1 - V_2 - U_2^*}{U_2^*}\right) + I_1\left(1 + \frac{V_1 - V_2 - U_2^*}{U_2^*}\right) \\
&= V_1 + \frac{V_1 - V_2}{U_2^*}\left(U_1^* + I_1\right) \\
T &= C + N \\
N &= T - C \\
&= T - V_1 - \frac{V_1 - V_2}{U_2^*}\left(U_1^* + I_1\right)
\end{aligned} \tag{S47}$$

So using these assumptions, we can estimate the coverage and the targeted fraction:

$$\begin{aligned}
L &= \frac{V}{C} = \frac{V_2}{V_1 + \frac{V_1 - V_2}{U_2^*}\left(U_1^* + I_1\right)} \\
p_{\text{in}} &= \frac{C}{T} = \frac{V_1 + \frac{V_1 - V_2}{U_2^*}\left(U_1^* + I_1\right)}{T}
\end{aligned} \tag{S48}$$

Both of these equations consist only of the measured values. Since the model is an approximation, there may be other effects, but these relations can provide a useful guide to the value of the study parameters that contribute to bias.

### S8 Alternative Scenario Translation

In the main text, we described applying this model to evaluating a novel vaccine during an Ebola outbreak, but noted in the discussion that the approach could be generically applicable. The previous sections outline the model in generic terms. Here we provide an example translation of that generalisation to another case: a vector control intervention for dengue.

In this scenario, we consider an intervention like indoor residual spraying, applied to urban households on a block basis (*i.e.* set of contiguous households, determined by street intersections) ahead of the dengue season. Some blocks would get no coverage (*i.e.* be amongst the non-targeted population), while others would receive coverage at some level (with non-coverage corresponding to *e.g.* availability to let treatment teams into house on that day or presence of children under some age).

Later, during the dengue season, people in the study population would seek healthcare with symptoms that would lead to testing for dengue, corresponding to the primary process. However, because dengue is frequently asymptomatic, the secondary process would be to test individuals in the primary cases household and adjacent households. Instead of a contacts-based secondary route, there is geospatial secondary route.

Whether a TND study would be ideal for this scenario is certainly a topic for debate. However, it is possible to frame this scenario and other potential pathogen spread and surveillance processes in the same terms we have introduced in this analysis.
